# A novel analytic framework to investigate differential effects of interventions to prevent obesity in children and young people

**DOI:** 10.1101/2024.03.07.24303614

**Authors:** F Spiga, AL Davies, JC Palmer, E Tomlinson, M Coleman, E Sheldrick, L Condon, THM Moore, DM Caldwell, FB Gillison, S Ijaz, JD Nobles, J Savović, R Campbell, CD Summerbell, JPT Higgins

## Abstract

**Background:** Recent systematic reviews and meta-analyses on the effects of interventions to prevent obesity in children aged 5 to 18 years identified over 200 randomized trials. Interventions targeting diet, activity (including physical activity and sedentary behaviours) and both diet and activity appear to have small but beneficial effects, on average. However, these effects varied between studies and might be explained by variation in characteristics of the interventions, for example by the extent to which the children enjoyed the intervention or whether they aim to modify behaviour through education or physical changes to the environment. Here we develop a novel analytic framework to identify key intervention characteristics considered likely to explain differential effects.

**Objectives:** To describe the development of the analytic framework, including the contribution from school-aged children, parents, teachers and other stakeholders, and to present the content of the finalized analytic framework and the results of the coding of the interventions.

**Design and methods:** We first conducted a literature review to find out from the existing literature what different types of characteristics of interventions we should be thinking about, and why. This information helped us to develop a comprehensive map (called a logic model) of these characteristics. We then used this logic model to develop a list of possible intervention characteristics. We held a series of workshops with children, parents, teachers and public health professionals to refine the list into a coding scheme. We then used this to code the characteristics of each intervention in all the trials which aimed to prevent obesity in children aged 5 to 18 years.

**Findings:** Our finalized analytic framework included 25 questions across 12 characteristics. These addressed aspects such as the setting of the intervention (e.g. at school, at home or in the community), mode of delivery (e.g. to individuals or to groups children), whether the intervention targeted diet and/or activity, complexity (e.g. focused on a single swap of juice for water or aimed to change all aspects the diet), intensity, flexibility, choice, mechanism of action (e.g. through participation, education, change in the social environment, change in the physical environment), resonance (e.g. credibility of the person delivering the intervention), commercial involvement and the ‘fun-factor’ (as perceived by children). We coded 255 interventions from 210 randomized trials.

**Conclusions:** Our evidence-based analytic framework, refined by consulting with stakeholders, allowed us to code 255 interventions aiming to prevent obesity in children aged 5 to 18 years. Our confidence in the validity of the framework and coding results is increased by our rigorous methods and, especially, the contribution of children at multiple stages.

**Funding:** This article presents independent research funded by the National Institute for Health and Care Research (NIHR) Public Health Research programme as award number 131572.

**Plain language summary:** More children and adolescents worldwide are developing overweight and obesity. Being overweight at a young age can cause health problems, and people may be affected psychologically and in their social life. Children and adolescents living with overweight are likely to stay that way or develop obesity as adults and continue to experience poor physical and mental health.

It is important to understand whether attempts to help children and young people modify their diet or activity levels (or both) reduce the chance that they develop obesity. In previous work we found that over 200 randomized trials have been done in people aged 5 to 18 years. These examine different strategies to try and prevent obesity. Whilst we found that these strategies have small beneficial effects on body mass index (BMI) *on average*, a notable finding was that there was a lot of variation in their results across the studies.

We want to understand what causes some strategies to be more effective than others. To do this we need to re-analyse the results of the studies. To inform this analysis, we developed a list of key characteristics that we and others thought would be likely to explain the variability in effects. We used this list to code over 250 strategies that have been studied. The development process included review of literature and patients/public involvement and engagement (PPIE) that is extensive consultation with children, young people, parents, schoolteachers and public health professionals. Our final list included features such whether the strategy was based at school or in the home, whether the strategy targeted diet or activity, how long and intense the strategy was and how flexibly it could be implemented. We also included the ‘fun-factor’ of engaging with the intervention, for which we invited children and young people to help us out with the coding.

## Background

Population levels of overweight and obesity in childhood are a significant global challenge (1). From 1990 to 2022, age-standardised prevalence of obesity increased in girls in 186 countries (93%) and in boys in 195 countries (98%); in most countries, obesity more than doubled (2). Children and adolescents living with obesity are more likely to experience reduced health-related quality of life and, for adolescents, a number of comorbidities including type 2 diabetes mellitus, fatty liver disease and depression (3).

We recently conducted two systematic reviews and meta-analyses of over 200 randomized trials of interventions aimed at preventing obesity in children and young people (CYP) aged 5-11 and 12-18 years, respectively (4, 5). Within each age group, we performed meta-analyses of body mass index (BMI), age-and sex-standardized BMI (zBMI) and BMI percentile results, comparing interventions targeting diet, activity (including physical activity and sedentary behaviour) or a combination of both. Our findings suggest that activity interventions, alone or in combination with dietary interventions, can have a modest beneficial effect on obesity. However, there was evidence of substantial statistical heterogeneity (i.e. that effects varied substantially from study to study) in 26 of 54 primary analyses. Prespecified subgroup analyses by main setting of the intervention (school, home, school and home, other), country income status (high vs non-high), participants’ socioeconomic status (low vs mixed) and duration of the intervention (short vs long; age group 5-11 studies only) did not sufficiently explain the heterogeneity among the studies.

This heterogeneity is likely to be due in part to variation among the interventions within each category (dietary, activity, and combined), since the interventions examined varied notably in nature, setting, complexity, delivery, intensity and duration. Variation in results will also arise from differences in the participants, and potentially because of different biases in the studies. These sources of heterogeneity not only present a statistical problem; they pose challenges for decision-making and for planning future studies. The work described in this paper arose from our desire to investigate the heterogeneity across the substantial body of evidence containing over 200 randomized trials. A protocol for the project was posted in advance on the funder’s website (https://fundingawards.nihr.ac.uk/award/NIHR131572).

We sought to develop a strategy for examining features of the interventions that might be associated with greater or lesser effectiveness. While public health guidance for developing obesity prevention initiatives is available (6) and taxonomy related to childhood obesity prevention has been developed to inform meta-analysis (7), our project required a bespoke scheme for coding each of the interventions studied.

Here we describe the development and coding of the analytic framework. Specific steps in the development of the framework included: review of existing logic models and analytic frameworks; refinement of our existing logic model; identification of key features of the interventions; contribution of CYP, schoolteachers and public health professionals; development and piloting of the final framework; and preparation of a coding manual.

We describe how we coded the interventions in collaboration with children and young people and how we analysed the data, and we report the results of the coding. In subsequent work to be described elsewhere, we will report the statistical methods developed specifically for the synthesis and the results of the application of these methods to the data coded according to the analytic framework. The analytic framework comprises a logic model to refer to the general characteristics that are relevant to the problem (which we illustrate graphically) and a coding scheme that pulls out components of the logic model to inform the synthesis.

## Methods

### Development of the analytic framework

Development of the analytic framework consisted of four phases (Figure 1): (i) drafting of a preliminary logic model, (ii) refinement of the logic model; (iii) consultation with CYP and their parents, our research advisory group (including academics expert in the field and two young people), teachers, and public health professionals; (iv) development of a coding scheme. We describe each phase below.

**Figure 1:**
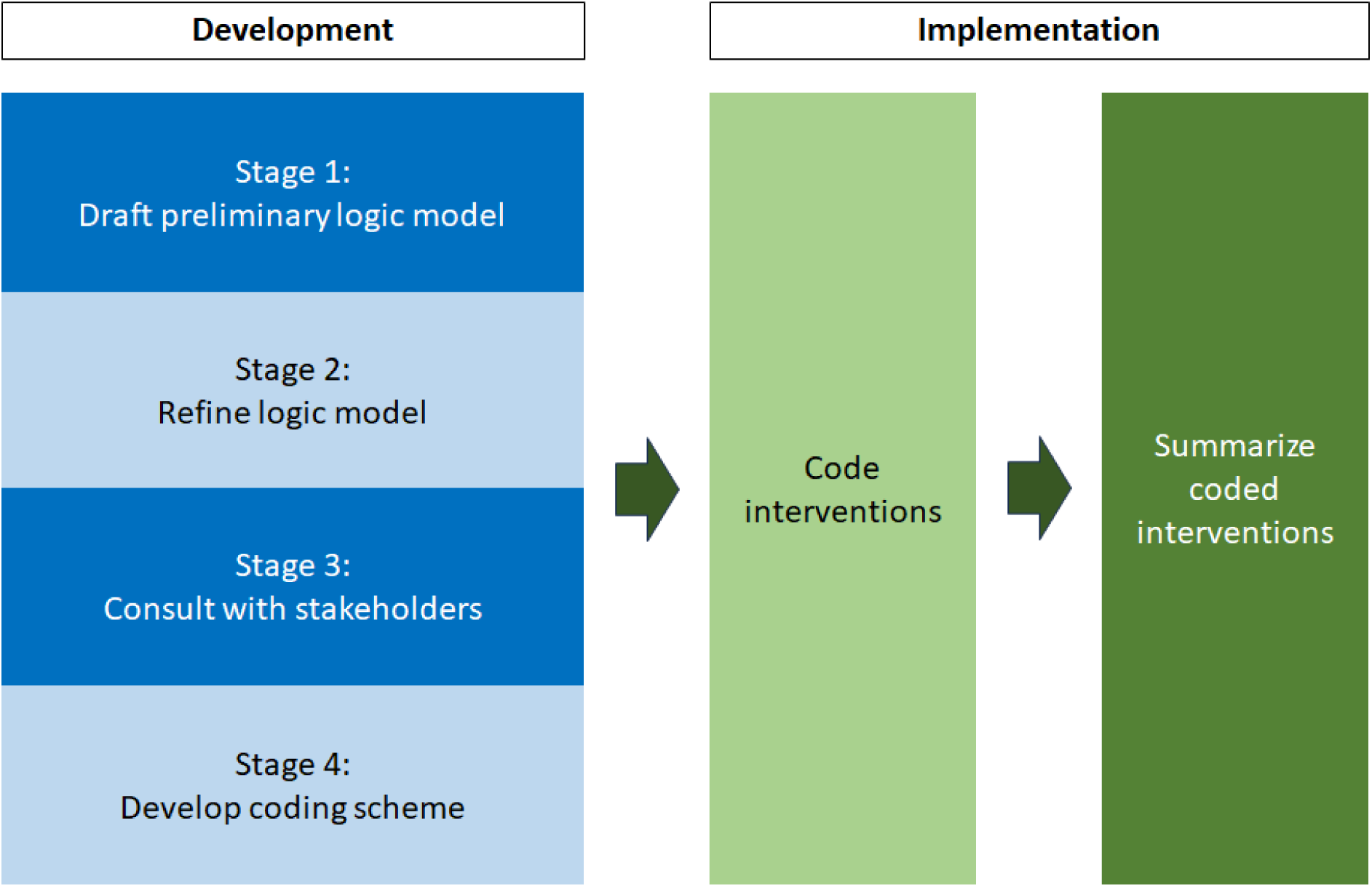
An overview of the stages of the development and implementation of the analytic framework.

#### Preliminary logic model (Figure 1, <u>Stage 1</u>)

Following advice from Cochrane (8) we drafted a preliminary logic model while drafting the review protocol (https://fundingawards.nihr.ac.uk/award/NIHR131572) to organize our initial thoughts on potentially important intervention and population features. Elements of the logic model included the type of intervention (9, 10), the setting (11) and the mode of delivery (12). Since our interventions of interest seek to change dietary and/or activity behaviour, the preliminary model also drew on elements from the COM-B behaviour change framework (13) and a complex adaptive systems perspective (14). We additionally drew on previous work in which we employed a ‘wider determinants of health’ (WDoH) perspective to characterize obesity interventions studied in obesity prevention trials in children, using a de novo ‘mapping tool’ developed to cover 226 potential causes of obesity (14). This analysis revealed that many of the studied interventions were aligned with the individual lifestyle factors domain of WdoH, many with the living and working conditions domain and some with social and community factors. In the light of this, we considered contextual factors that may influence BMI. We also drew on our realist review addressing the contextual and mechanistic factors associated with successful interventions in schools (15).

The preliminary logic model (see Appendix 1) included the concepts of setting (e.g. school, home, region, country); participant characteristics (e.g. age, sex, socioeconomic status); intervention characteristics including function (e.g. education, training, enablement); the targeted behaviour (e.g. diet, activity); intensity, sources of behaviour change (motivation, capability, opportunity); how it is experienced by the child (e.g. one-to-one, group based); who is targeted by the intervention (e.g. child, parent, community); and who delivers the intervention (e.g. self-delivered, parents, teachers). The logic model also included short term outcomes (e.g. changes in social and physical environment, empowerment of providers and/or children/families), medium term outcomes (e.g. improved diet and physical activity) and our target long-term outcome of reduced incidence of obesity.

#### Refinement of the logic model (Figure 1, <u>Stage 2</u>)

To formalize and refine our preliminary logic model after the project was funded, we undertook a scoping review of existing logic models and analytic frameworks in the fields of (i) obesity prevention, (ii) behavioural change and (iii) assessment of complex interventions in the context of systematic reviews. We searched PubMed using phrases such as “analytic framework and obesity”, “logic model and obesity”, “analytic framework and behavioural change”, “logic model and behavioural change”, “complex interventions”, examined reference lists and consulted with collaborators. Our search was not intended to be systematic, since we aimed to identify a wide, rather than a comprehensive, selection of ideas to refine our logic model.

#### Stakeholder consultation (Figure 1, <u>Stage 3</u>)

The third stage of development of the analytic framework was to share the list of intervention features from the logic model with stakeholders. We sought input on what intervention features or components could be used to characterize the available studies and lead to building blocks of future interventions or their implementation. As per the Health Research Authority/NIHR INVOLVE statement (16), ethical approval was not required for the contribution of the public as part of the patient and public involvement and engagement.

##### Children, young people and teachers

We took the view that children and young people have much to contribute to the design and delivery of interventions targeted at them, particularly when processes that respond to their preferences for engagement support them to share their views (17). We therefore started with two workshops to engage this audience, the first with a group of five CYP aged 12-18 years on their own, and the second with a group of six CYP aged 12-16 accompanied by a parent (January 2022). We identified participants through Bristol’s Generation R Young People’s Advisory Group (YPAG; https://generationr.org.uk/bristol/), a group funded by the National Institute for Health and Care Research (NIHR) Applied Research Collaborations (ARC) West and the Bristol Biomedical Research Centre (BRC). YPAG comprises CYP aged 10 to 22 years who are interested in healthcare and research, offering an opportunity for them to evaluate critically the way research about them takes place. Both workshops were also attended by one of the YPAG coordinators, who chaired the meeting, and by four members of the project team. We later held two online meetings with teachers and head-teachers (January 2023) which were attended by three and five teachers, respectively.

The approach we used to elicit input was similar in all four of these workshops. We started the meeting by asking the group “What should we do to prevent childhood obesity?” We then provided some examples of interventions we have included in our reviews and asked some more specific questions (see Box 1).

###### Box 1. Questions asked at workshops with children, young people, parents and teachers

1. What should we do to prevent childhood obesity?
2. What sorts of approaches do you think might work?
3. From the ideas generated, what sorts of approaches might work best?
4. Are there approaches that might work well across all age groups? Or that might differ importantly?
5. Are there approaches that might work particularly well for those most likely to gain weight?
6. Are there combinations that might be particularly good or particularly bad?

##### Public health professionals

We held an online meeting with public health professionals from local authorities in our region (January 2023), which was attended by five public health experts, one young person member of our advisory group (see below), one schoolteacher from our workshop with teachers and six project staff. After a brief introduction to the project and presentation of the latest list of important intervention features, we discussed each item and specifically asked for feedback on the relevance of features included; whether there were important features missing; and whether any should be dropped. We also discussed whether any of the features may work better in tandem (i.e. have interaction or synergistic effects). In addition to this meeting, we held one-to-one (or two-to-one) meetings with various public health professionals who could not attend the group meeting.

##### Advisory group

Alongside these, we consulted with our project advisory group. Our advisory group comprised international academics with expertise in the field and two young people aged 15 and 16 years. We presented a preliminary version of list of intervention features at our annual advisory group meeting in February 2022 (online), attended by six advisory group members, one YPAG coordinator and four project staff. We started the meeting by outlining the themes emerging from the workshops with the children and discussed these themes with the aim of reducing these ideas to a smaller number of generic codable features. The specific discussion points were (i) to consider whether it was feasible to code the items currently included; (ii) to consider how to code the items; and (iii) to identify any additional concepts.

#### Development of coding scheme interventions (Figure 1, Stage 4)

We created a coding scheme out of the final list of intervention features. Since the coding scheme would feed directly into the statistical analysis, we established the following informal criteria for the scheme so as to maximize our prospect of obtaining informative results: (i) each item in the coding scheme should be applicable to every intervention examined in the studies; (ii) each item should ideally be a dichotomous variable that approximately divides the studies into halves (since this would maximize precision in the estimation of the regression coefficients); (iii) the coding scheme should include as many intervention features that potentially impact on effectiveness as possible (iv) the number of items should be kept to a minimum. There is clearly a tension between the last two criteria. To try and meet (iii), we considered all the features identified by stakeholders. To try and meet (iv), we bore in mind that rules of thumb generally advocate at least 10 data points per predictor in regression analyses, suggesting that at most 25 items should be included.

The questions in the coding scheme were formulated to elicit binary responses (‘Yes’/’No’) or using a very small number of categories, for the purpose of inclusion in our statistical model. There were two exceptions, both relating to intervention duration for which responses were collected in number of weeks.

In addition to features of the interventions, we added to the coding scheme some features of the trial participants that might impact on intervention effectiveness: age group, income category of the country in which the trial was performed and whether the trial specifically targeted individuals of low socio-economic status.

We wrote a guidance document to explain each of the items in the coding scheme.

### Implementation of the coding scheme

#### Data set

The set of trials to which we applied the coding was derived from two Cochrane Reviews of interventions to prevent obesity in children aged 5-11 and in CYP aged 12-18 (4, 5). We coded only studies that were included in meta-analyses in these reviews and therefore had valid data for inclusion in the planned complex synthesis (to be reported elsewhere). Because intervention coding was conducted at intervention level and not at study level, for each study we had to consider (i) whether the reference arm was a control group such as no intervention or ‘usual care’, or an eligible active intervention (i.e. the trial made a ‘head-to-head’ comparison); and (ii) whether more than one intervention was implemented in each study (i.e. the trial was a multi-arm study). We coded only active interventions in controlled trials and coded all active interventions in multi-arm studies.

#### Piloting

We piloted the coding in several waves. One reviewer (FS) first tested the framework on five studies. Two reviewers (FS and ALD) then independently piloted the framework on ten studies that were purposefully selected to provide a diverse collection. In the third wave a further twenty studies were coded by two different pairs of reviewers. (FS and ALD, FS and JCP). After each wave of the piloting, we recorded and discussed issues identified the among the project team and implemented appropriate modifications to the coding scheme and/or coding manual as necessary to achieve consistent and comprehensive capture of study features, following previous methods (18).

#### Coding

Following the piloting phase, we used the finalized coding scheme for application to the interventions described in the remaining studies. Two reviewers (from FS, ALD, JCP, ET, THMM, DMC and JPTH) independently coded each study using the data extracted during the Cochrane Reviews, with recourse to the full study reports as necessary. All coding discrepancies were resolved by discussion and in case of disagreement a third reviewer was involved.

##### Contribution of children and young people

One of the intervention features that emerged from talking with the CYP was the importance of the intervention being enjoyable (to use their words, having the ‘fun-factor’). Inspired by a discussion with the children we decided that the most appropriate people to code an item about this would be CYP themselves. We recruited a panel of young people from the Bristol YPAG by emailing the group with an explanation of the aim of the project and the task involved. We supplemented volunteers from the group with younger children known to members of the research team.

From each study, we extracted a brief description of the intervention(s). We compiled these into batches of ten intervention strategies. For each intervention, the documentation included a strategy ID (study name and year), the intended age group (i.e. the mean age or age range of the target children as reported in the study) and the setting of the interventions (see Appendix 2). We asked the CYP to read the description of each intervention and then answer the following two questions using an online survey via *Online surveys* (https://sscm.onlinesurveyfigures.ac.uk), with possible answers being: ‘really boring’/’a bit boring’/’neutral’/’a bit fun’/’really fun’:

- Question 1: How enticing would **you** find this strategy?
- Question 2: How enticing do you think **children in the intended age group** would find this strategy?

Our primary interest was in Question 2. Question 1 was included for us to learn about the interests of our volunteers and in the hope that it would reduce the impact of personal preferences on their answer to Questions 2. We also gave the CYP the opportunity to comment on the specific interventions by providing an optional free-text box (Figure 2).

**Figure 2:**
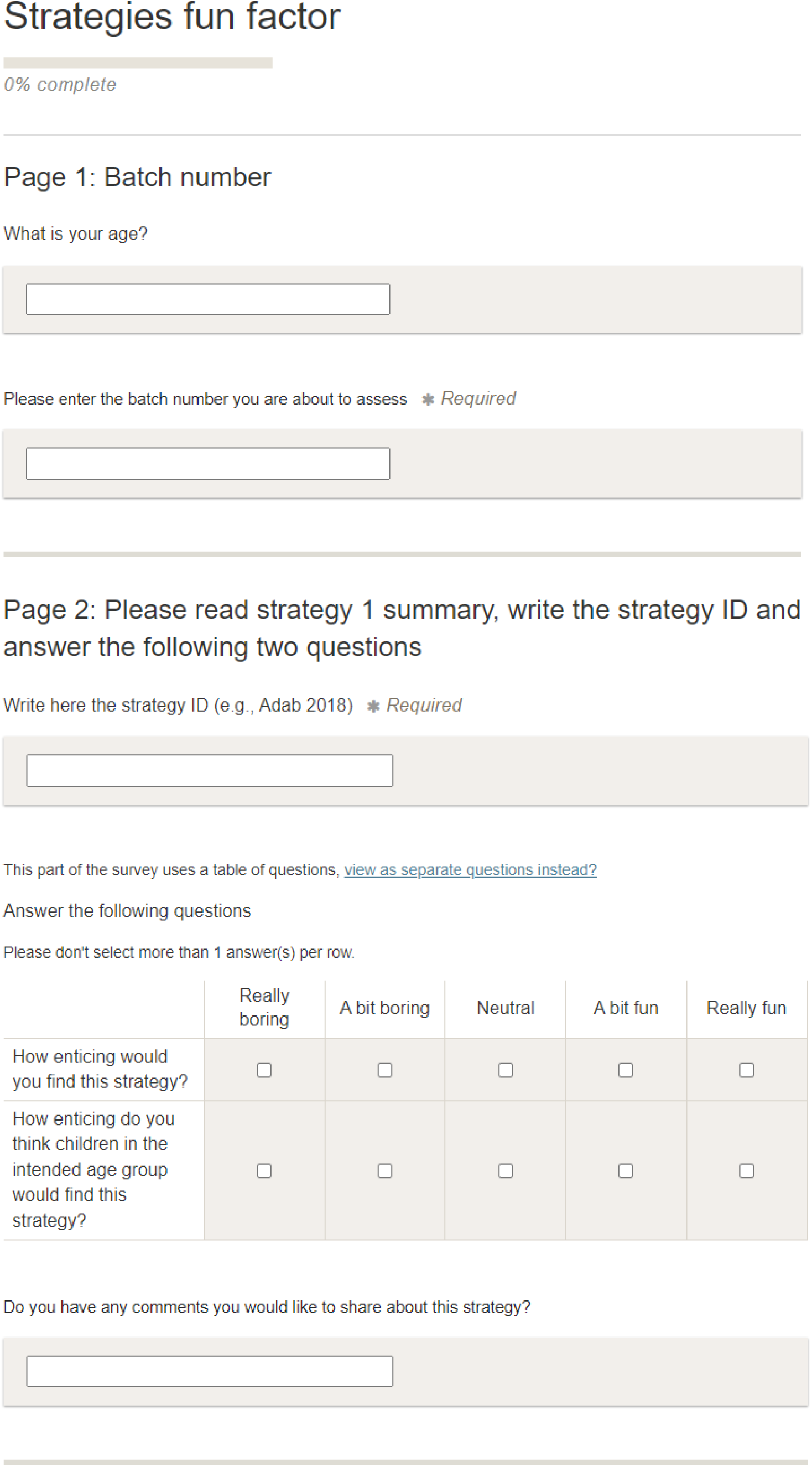
Fun factor coding survey.

The volunteering CYP decided how many batches of the interventions they wanted to assess. We ensured that each intervention was coded by at least four CYP. In case of multiple participating CYP from the same household, we assigned a different batch of interventions to each. We compensated the volunteers £25 for the completion of each batch of ten interventions. We did not develop a strategy for resolving discrepancies; instead, we developed an algorithm to determine a judgement based on the individual responses, described in the following section.

#### Analysis of the coded data

We analysed the coded data for all the active intervention arms separately for the two age groups 5-11 years and 12-18 years. For each item with categorical responses (e.g. ‘Yes’/’No’) we calculated the number of interventions falling into each possible category and expressed these as percentages. We converted total and peak duration into binary variables (short or long) by dichotomizing at the medians of the reported values. For these, we also present the means and standard deviations (SDs) of the quantitative data.

For the fun factor, we had four distinct responses from four volunteer CYP. We first combined the ‘really fun’ and ‘a bit fun’ categories and combined the ‘really boring’ and ‘a bit boring’ categories. We then classified an intervention as ‘fun’ if, across the four (or more) coders, either the majority of coders regarded it as fun or an equal number of coders regarded it as fun and neutral. We classified an intervention as neutral if equal numbers of coders regarded it as fun and boring. We classified an intervention as boring otherwise (i.e. if either the majority of coders regarded it as boring, or an equal number of coders regarded it as boring and neutral). We refer to this approach as category-based analysis for consensus fun factor (CACFF). We performed a sensitivity analysis in which we calculated the numerical average response by assigning the following values to each possible answer given by each coder for each intervention: really boring = 1, a bit boring = 2; neutral = 3; a bit fun = 4; really fun = 5. We then classified each intervention as fun (mean >3), neutral (mean = 3) or boring (mean <3). We refer to this method as number-based analysis for consensus fun factor (NACFF).

## Results

### Refined logic model

Our informal scoping review of other models and frameworks identified relevant academic papers in the field of obesity prevention (19–25) and related fields in which interventions aimed at behaviour change were described (26–28). Our scoping also identified guidelines to assess complex interventions in systematic reviews (29–31) and frameworks that address equity in the context of evidence synthesis, including the PROGRESS-PLUS framework (32). In order to translate key aspects of the interventions (e.g. complexity) into questions, we referred to published guidelines including (33–35). These additional frameworks gave us insights into further characteristics of the intervention that are likely to be important for their effectiveness such as the target, the complexity of the interventions, and the role of the community. Guided by the PROGRESS-PLUS framework we also implemented a more comprehensive description of the participants characteristics.

The refined version of our logic model is available in Appendix 3. The preliminary logic model was modified to expand participant characteristics (including the PROGRESS-PLUS framework (32)); we also made substantial modifications to the intervention characteristics to include duration; complexity (e.g. simple or multiple components); fidelity (i.e. whether the intervention was implemented as intended); whose behaviour the intervention aims to change (e.g. child, parent, community), and other characteristics (e.g. participation, flexibility) We did not implement any changes in the setting and outcomes.

### Feedback from consultation

Children, young people and their parents emphasized the importance of (i) thinking differently about different age groups (primary vs secondary school age); (ii) infrastructural changes (e.g. improved dining facilities in schools), (iii) engaging families in achieving behavioural change; and (iv) if those delivering the intervention had credibility with or were role models for the CYP. Additional important features of the intervention that emerged from talking with the young people were (v) the adaptability and flexibility of the intervention (e.g. children should be able to choose their favourite sporting activity); and (vi) the importance of the intervention being fun. A full list of themes that we addressed is reported in Appendix 4. Consultation with children, young people and their parents led to a list that comprised 17 categories including: realm targeted, multifactor-ness, intensity and duration, theory, mechanism of action (i.e. change children’s dietary or activity behaviour by making them do something, educating them or changing their social and/or physical environment), commercial interests, integration, choice, fun factor, messaging (i.e. how the intervention is ‘sold’ to the children/young people), resonance, peer support, community engagement, setting, recipient, targeting and fidelity.

Teachers commented on the setting for different types of interventions: for example, physical activity interventions are readily delivered at school, whereas it is more difficult to control children’s diets if they bring lunch boxes from home. They also discussed the importance of role models and whether teachers are the most appropriate to provide guidance. They mentioned resource and time constraints, and that embedding the programme within the curriculum may be more efficient than changing the existing curriculum. They thought that it was important for the intervention to be sustainable in the long-term. Furthermore, they highlighted the importance of involving the parents to ensure continuity of school-based interventions (e.g. school-based cooking classes followed by meal boxes delivered at home for children and parents to prepare the meals together). Our discussions with the teachers also highlighted the importance of empowering the children (e.g. involving them in preparing home meals) and considerations for the different age groups (e.g. educational interventions may be more effective in younger children, because older children are more independent). The teachers also suggested that it may be effective to link interventions to mental health outcomes as these are of paramount interest to young people these days.

Discussions with our project advisory group resulted in some features being dropped from our list of intervention features. Some items were judged to be less informative than others (e.g. whether the intervention was theory-based); others as overlapping with other components (e.g. who was targeted by the intervention, whether there was community engagement); and others as unfeasible to code due to lack of information (e.g. fidelity in implementation of the intervention). Items recommended to be retained as important included consideration of who is delivering the intervention (and in particular the resonance it would have with the children), seeking to influence the child’s social environment as part of the mechanism of action, and the complexity of the interventions (e.g. in terms of how many dimensions or factors it comprises). Crucially, it was advised that it is “*important to code the things that are important to the young people and their parents*”.

Our final discussions with public health professionals reinforced many of the points mentioned above and helped us refine the list of items substantially. Although no additional components were included at this stage, some questions and answers were reworded for sake of, precision, clarity and unambiguity; for example, in the item about children’s choice in how they modified their diet or activity, the question ‘Is there choice of activity/diet within the intervention?’ was amended to ‘Is choice of activity/diet designed into the intervention?’ and the realm targeted item the answer ‘Yes’/’No’ was amended to ‘Yes exclusively or substantially’/’Yes minimally’/’No’.

### The finalized coding scheme and coding manual

The finalized version of the coding scheme comprises 25 individual questions spread across 12 categories (Table 1). We largely achieved our aim of formulating a series of questions that can be answered for each intervention with a small number of possible answers. The coding manual contains, for each item, a detailed explanation of the question and a selection of examples illustrating how the interventions should be coded. A copy of the manual is available in Appendix 5. Below we provide a brief explanation of each of the 12 categories with examples.

**Table 1.** Finalized analytic framework.

| Item | Question (*possible answers*) |
| --- | --- |
| **1. Setting** | Is the intervention delivered in a school (in full or in part)? (Yes/No) |
|  | Is the intervention delivered in the home (in full or in part)? (Yes/No) |
|  | Is the intervention delivered in the community or other non-school and non-home setting (in full or in part)? (Yes/No) |
|  | Does the intervention include a home activity? (Yes/No) |
| **2. Mode of delivery to the child** | How is the intervention delivered? (*Exclusively or mainly individually/Both individually and as a group/Exclusively or mainly as a group*) |
|  | Is the intervention delivered electronically? (*Yes exclusively/Yes significantly/Yes as a minor component/No*) |
| **3. Realm targeted** | Does the intervention aim to change diet? (*Yes exclusively or substantially/Yes minimally/No*) |
|  | Does the intervention aim to change activity levels? (*Yes exclusively or substantially/Yes minimally/No*) |
| **4. Multifactor-ness & Dimensionality** | Does the intervention use multiple strategies (three or more)? (Yes/No) |
|  | Is the intervention applied in a single phase? (Yes/No) |
|  | Is the intervention applied for a continued period? (Yes/No) |
| **5. Peak intensity and duration** | During how many weeks does the whole intervention last? (*Numerical; to be dichotomized at the median*) |
|  | For how many weeks does the peak engagement period of intervention last? (*Numerical; to be dichotomized at the median*) |
|  | What is the level of engagement with the children? (*High/Low*) |
| **6. Integration** | Is the intervention integrated into the normal curriculum/ habits? (Yes/Partially/No) |
| **7. Flexibility** | Is the intervention designed to be implemented in a flexible manner/tailored to specific participants? (Yes/No) |
| **8. Choice** | Is choice of activity/diet designed into the intervention? (Yes/No) |
| **9. Fun Factor** | How enticing would you find this strategy? (*Boring/Neutral/Fun*) |
|  | How enticing do you think children in the intended age group would find this strategy? (*Boring/Neutral/Fun*) |
| **10. Resonance** | Is the intervention experienced by children via someone external or unusual? (Yes/No) |
| **11. Mechanism of action and recipient** | Does the intervention have an explicit component that requires the child to participate? (Yes/No) |
|  | Does the intervention have an explicit component of education/information provision for the child? (Yes/No) |
|  | Does the intervention have an explicit component aiming to change the social environment of the child? (Yes/No) |
|  | Does the intervention have an explicit component aiming to change the physical environment of the child? (Yes/No) |
| **12. Commercial interests** | Are commercial interests involved in the trial and/or intervention? (Yes/No) |

### Coding categories

#### Setting

This is a measure of the setting where the intervention is delivered. Possible answers were ‘school’, ‘home’ or ‘community or other non-school/home’ (e.g. club, gym, shop, library, healthcare centres). Within *setting* we also coded each intervention according to whether the intervention protocol included home-based activities for the children (e.g. cooking or games activities with parents, additional homework).

#### Mode of delivery to the child

This is a measure of how the child experiences the intervention, that is, as an individualized intervention (e.g. a leaflet about healthy meals given to each student at school; a visit to an healthcare centre, homework with parents, a website to view at home), through a group of children (e.g. school classes or scout troop meeting), or both (e.g. school classes and homework activities). Within *mode of delivery,* we also coded the intervention according to whether it was delivered electronically (i.e. via digital media, online website or app) and in what capacity (i.e. exclusively, significantly, as a minor component or not at all).

#### Realm targeted

This is a measure of whether the intervention seeks to change ‘diet’ (e.g. introduction or replacement of food beverages with healthier options; re-organization of food display in the school canteen or in shops; education on healthy diet; cooking classes; healthy meal box for the family), ‘activity’, including increase in physical activity (e.g. modified or additional physical activity classes at school) and/or reduce sedentary time at home (e.g. active video games), or ‘both diet and activity’, and in what capacity (i.e. exclusively or substantially to indicate the main component, minimally to indicate a minor component, or not at all).

#### Multifactorness /dimensionality

This is a measure of how complex the intervention is, including how many ways the children are targeted, e.g. at multiple levels or in multiple phases. Questions within this category include whether the intervention has multiple components, that is, uses at least three different strategies (e.g. classroom activities, changes in the canteen food and homework activities), is delivered in multiple phases, that is, uses different strategies or settings at different times (e.g. a more active phase followed by a less active “maintenance” phase or a “top-up” phase), and is delivered in a continuous manner, that is without breaks between the beginning and the end of the intervention (during the whole school-year) or for a discontinuous period (e.g. lectures delivered for 12 weeks/year for two years).

#### Peak intensity and duration

This is a measure of how intensely the intervention is experienced by the child and it covers the duration and frequency of the intervention. Questions within this category cover the duration in weeks of the whole intervention and of the peak engagement (if different from the whole intervention). The category also measures the level of engagement with the children during the peak period, using the number of sessions of engagement per week as guidance so that the intervention are coded as ‘high’ engagement if there was at least one session of engagement with the children per week and ‘low’ if there was less than one session of engagement with the children per week.

#### Integration

This is a measure of the extent to which the intervention is ‘normalized’ within the school curriculum or normal habits of the child (e.g. as part of regular homework). This measure provides an indication of how much ‘extra effort’ (by the provider and/or the recipient) would be required for the intervention to be successful. Examples of interventions that are completely integrated include modification of physical activity classes or the addition or replacement of regular school meals with healthier options. Examples of interventions that are partially integrated are those with a combination of integrated activities and something extra (e.g. after school program or homework). Examples of interventions that are not integrated at all are those in which the school needs to add something to an existing programme (e.g. an extra physical activity class extending school hours or home activities with the parents) or when the child needs to sign up for/agree to after-school classes.

#### Flexibility

This is a measure of the extent to which the intervention can be implemented flexibly, within the intervention protocol. That is, whether an intervention is adapted to the particular classroom/household at teachers/parents’ discretion (e.g. an intervention consisting of the replacement of regular meals with healthy meals where the healthy meals are decided by each participating school kitchen staff).

#### Choice

This is a measure of the extent to which children are free to make the intervention work for them (e.g. an intervention in which the child is able to choose which sport they do, or which food to eat).

#### Fun factor

This is a measure of the extent to which the intervention is expected to be enjoyable for the age group to whom it is delivered. We anticipated that some interventions that involve games, songs, plays may look fun to everyone, whereas interventions that includes sport activities or cooking with the parents may not look fun to everyone and interventions that included classroom lectures or replacement of sugar sweetened drinks with water may not look fun to anyone. We also considered that some interventions may be appropriate for children aged 5-11 year but not for older children (e.g. a song about healthy eating), and *vice versa*, a video game intervention designed for older children (12-18 years old) may not be fun for a 5 years old child. We designed the questions and answers for this category to be suitable and appropriate for CYP as they were invited to help us with coding the interventions for this item (see methods section on fun factor).

#### Resonance

This is a measure of the extent to which the intervention is likely to attract the respect of the young people, particularly through the credibility of the person delivering the intervention. For example, an intervention may be experienced by children via someone external or unusual (e.g. a sport coach, a professional athlete, an influencer, a dietitian or a nurse) or someone familiar to them (form teacher or a parent/carer).

#### Mechanism of action and recipient

This is a measure of who is the direct recipient of the intervention (e.g. child, the teacher(s), parent(s), the child’s environment or others) and how the intervention aims to achieve a change in the child’s dietary and/or activity behaviour. Options for the latter are ‘participation’, ‘education’, ‘social environment’ and ‘physical environment’. An intervention that has an explicit component of modifying the child’s behaviour through participation is an intervention in which the child learns by doing something (e.g. a session of physical activity or a workshop on healthy nutrition in which the children are involved in cooking a meal). An example of an intervention that has an explicit component of education or information is the provision of literature or lessons in which there is no activity involving the child doing something. An example of an intervention that has an explicit component aiming to change the social environment of the child at school or home is an intervention in which teachers are instructed to encourage children to change their dietary or activity behaviours or parents are educated on healthy food. Examples of interventions that have an explicit component aiming to change the physical environment of the child at school or home are interventions that include placement of healthy foods in the school canteen, provision of exercise equipment at school or in the community, drawing running tracks in the playground or changing the school meal menu. For interventions using multiple mechanisms, we answered ‘Yes’ to all relevant options.

#### Commercial interests

This is a measure of whether commercial interests are involved in the trial or in the delivery of the intervention, such as an intervention within a study that was funded by industry (e.g. food or pharmaceutical industry) or an intervention that include use of equipment supplied by a manufacturer of sport equipment, or provision of food/drinks by a food supplier.

### Results of the coding

We coded 255 interventions from 210 randomized trials. Descriptive statistics summarizing the coding of these intervention arms are reported in Table 2. Results by age group are reported in Appendix 6 and the full data set is available in Appendix 7.

**Table 2.**
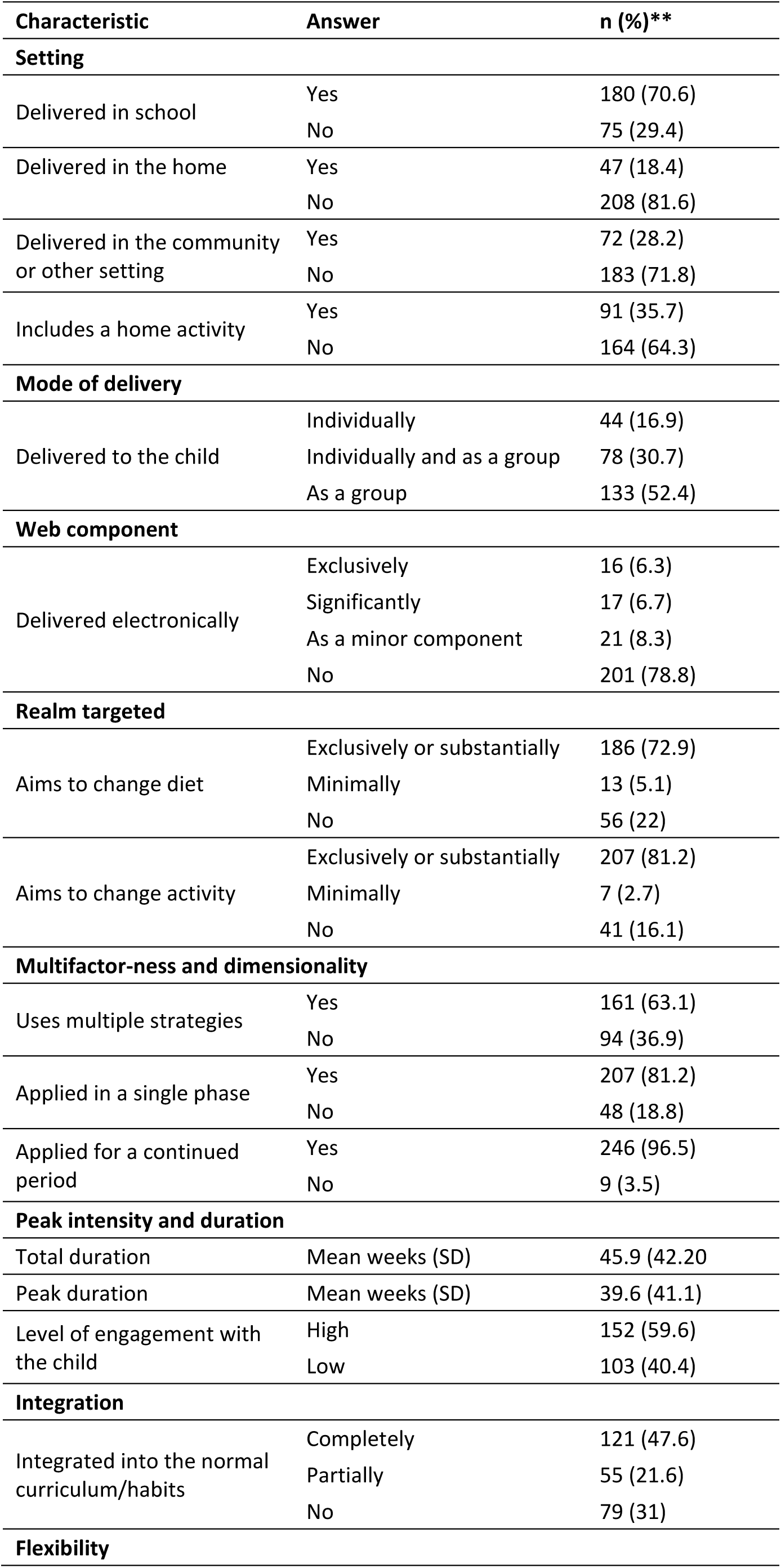
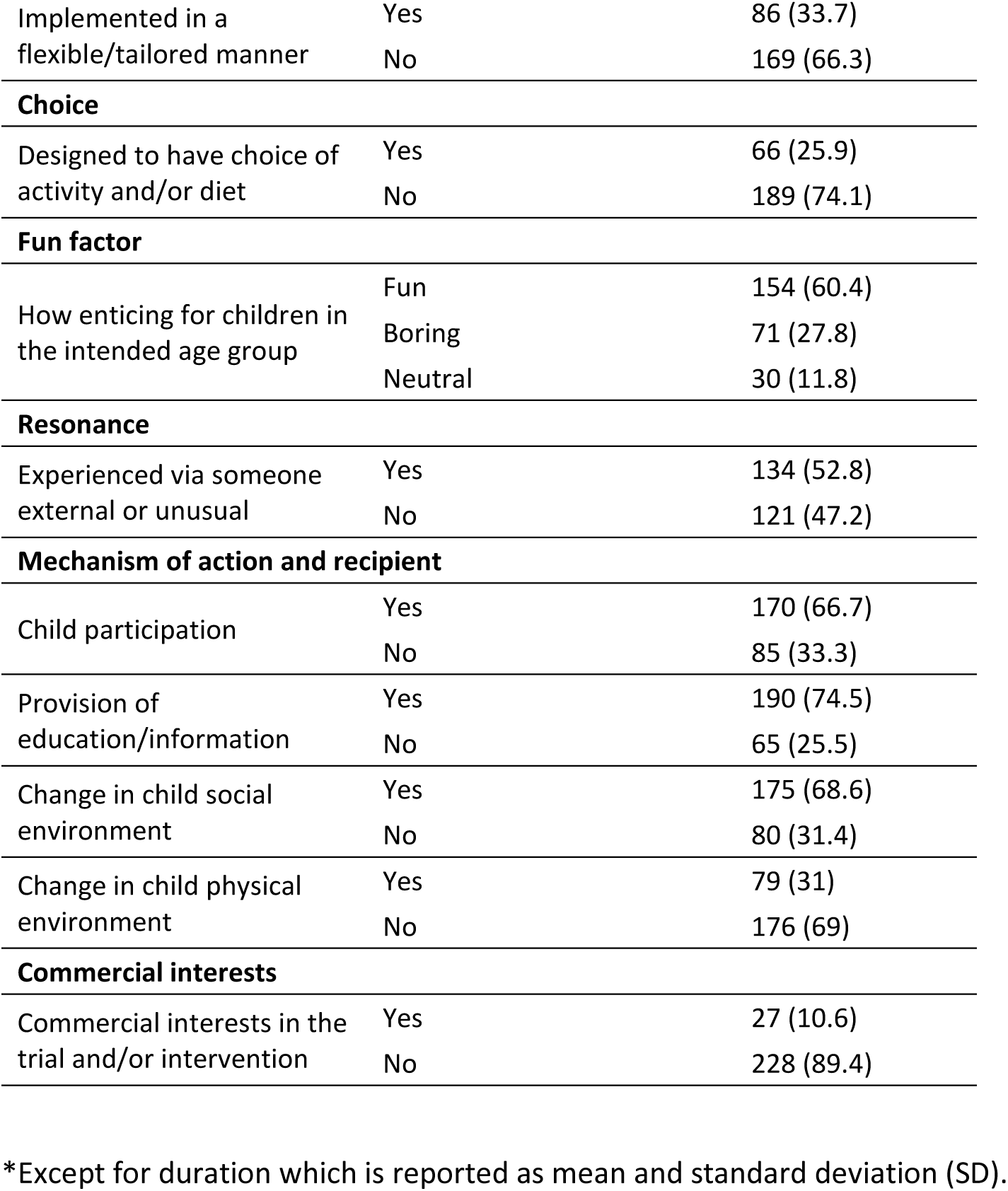
Coding results of all active intervention arms (n=255)

Of the 255 active intervention arms coded, 180 (70.6%) were delivered at school, 47 (18.4%) were delivered in the home and 72 (28.2%) were delivered in the community or other settings (e.g. primary care setting); 91 interventions (35.7%) included a home activity. Forty-four of the interventions were delivered individually (16.9%), 133 as a group (52.4%), and 78 (30.7%) were delivered both individually and as a group. Sixteen interventions (6.3%) were delivered exclusively electronically, 17 interventions (6.7%) included a significant electronic component and 21 (8.3%) included a minor electronic component. 186 interventions (72.9%) were aimed at changing diet exclusively or substantially and in 13 (5.1%) the component aimed at changing diet was minimal. There were 207 (81.2%) interventions aimed at changing activity (including increasing physical activity and reducing sedentary behaviour) exclusively or substantially and in 7 (2.7%) of these the component aimed at changing activity was minimal.

At least three different intervention components (or different strategies) were implemented in 161 interventions (63.1%), 207 interventions (81.2%) were applied in a single phase and 246 (96.5%) were applied for a continued period. The total mean duration of the intervention was 45.9 weeks (SD 42.2) with a mean peak period duration of 39.6 weeks (SD 41.1). The level of engagement with the children was high in 152 interventions (59.6%) and low in 103 (40.4%). The interventions were completely integrated in the normal curriculum or habits in 121 interventions (47.6%) and partially integrated in 55 interventions (21.6%). Eighty-six interventions (33.7%) were implemented in a flexible or tailored way and in 66 (25.9) there was an element of choice of diet and/or activity for the children. One-hundred-and-thirty-four interventions (52.8%) were delivered (partially or exclusively) by someone external or unusual. With regards to the mechanisms by which the interventions aimed at preventing obesity, 170 interventions (66.7%) required the child participation, 190 (74.5%) provided education or information, 175 (68.6%) changed the social environment of the child, and 79 (31%) changed the physical environment of the child. Commercial interests in the trial and/or intervention were found in 27 interventions (10.6%).

We received an overwhelming response from YPAG members to code the ‘fun factor’, with 31/89 CYP aged 12 to 18-years volunteering to participate in the project. Additionally, we recruited four children aged 6 to 13-years through colleagues at the University of Bristol. The 35 participants contributing to coding this item therefore ranged from 6 to 18-years of age. Among the 31 CYP who declared their ethnicity, one was Asian/Black/White, one Black African, one Indian, six Somali, one South Asian and 20 White British; 51% were female. According to our CACFF approach, 154 interventions (60.6%) were regarded as fun, 71 (27.8%) were regarded as boring, and 30 (11.8%) elicited neutral views (see Table 3). In our sensitivity analysis using the NACFF approach, we found discrepancy with the CACFF approach in just 9% of the interventions. When asked about their own views of the interventions (rather than the views of age-appropriate children in general), views were slightly more neutral, with slightly fewer being categorized as fun and slightly fewer as boring. We present examples of feedback received from the CYP on their experience of undertaking coding in Box 2.

**Table 3.** Coding results for fun factor (n=255)

| Categorical analysis for consensus of fun factor (CACFF) approach |  |  |  |  |  |
| --- | --- | --- | --- | --- | --- |
|  |  | ***Q2: How enticing do you think children in the intended age group would find this strategy?*** |  |  |  |
|  |  | **Fun** | **Neutral** | **Boring** | **Total** |
| ***Q1: How enticing would you find this strategy?*** | **Fun** | 124 | 9 | 13 | 146 |
|  | **Neutral** | 19 | 13 | 17 | 49 |
|  | **Boring** | 11 | 8 | 41 | 59 |
|  | **Total** | 154 | 30 | 71 | 255 |
| Sensitivity analysis: Numerical analysis for consensus of fun factor (NACFF) approach |  |  |  |  |  |
|  |  | ***Q2: How enticing do you think children in the intended age group would find this strategy?*** |  |  |  |
|  |  | **Fun** | **Neutral** | **Boring** | **Total** |
| ***Q1: How enticing would you find this strategy?*** | **Fun** | 130 | 11 | 13 | 154 |
|  | **Neutral** | 14 | 4 | 12 | 30 |
|  | **Boring** | 14 | 4 | 53 | 70 |
|  | **Total** | 158 | 19 | 78 | 255 |

##### Box 2. Feedback from children and young people on their coding of the ‘fun factor’

Alongside the ‘fun factor’ questions we also provided the coder with the opportunity to comment on the specific interventions. Some examples of feedback on interventions that they coded as fun were:

> *‘This strategy sounds very fun, integrating video games into it is a very good idea and will work extremely well’*.

> *‘I think this strategy is very good as it will involve education and skills as well as physical exercise.’*

> *‘Cooking classes for families and taster foods, games and tasting and cooking sessions with family members.’*

On interventions that the children coded as boring:

> *‘A bit too academic, could be taught in a more fun way.’*

> *‘I think incorporating normal school curriculum lessons with physical activity could take the fun out of it for some students.’*

> *‘I don’t think students this age would find lectures and doing group presentations to a class at all enjoyable.’*

Finally, on interventions that the children coded as neutral:

> *‘Kids may be reluctant to take advice from parents.’*

> *‘I think this strategy would be very effective but may be less interesting than others.’*

> *‘I think 10-year-olds will work well with their family and I like the idea of trying new recipes, but I think 10 sessions a month could feel like a lot.’*

We gave participants the opportunity at the end of their coding assignment to provide feedback on their experience. We received feedback from 14 participants (or their parents). Most of the feedback highlighted positive aspects of the project/task:

> *‘Thank you for this awesome opportunity it was great fun!’ (YPAG member)*

> *‘The process was amazing thanks for asking.’ (YPAG member)*

> *‘I think the process for this YPAG was very good and enjoyable to give feedback on.’ (YPAG member)*

> *‘I think as an activity it worked really well, no issues at all with the forms as I guess sometimes it’s difficult to fill out the same one twice with the same device/account so credits to that platform for allowing that for a task like this, maybe one to remember for next time. From my end it all seems well organized, documents were clear and not too complicated, forms were straightforward, and appreciated the extra box for additional comments if it was sometimes relevant.’ (YPAG member)*

We also received some valuable advice on how the coding process could be improved:

> *‘On the surface, I think the boys assumed the last piece of work would be easier than a meeting, but it proved more difficult as they found it a bit repetitive [….] the strategies were so similar that it was hard for them to come up with original comments…. Also, our youngest needed input from (older sibling) so he could understand the strategies.’ (Parent of YPAG child)*.

> *‘Overall, I think the form is quite straightforward to fill in. From the information I was given before doing the batches it sounded a bit complicated (in terms of different batches), however actually filling it in was relatively easy. I would say that having the feedback form and information on the same page would make the process easier because it got a bit confusing going back and forth through different tabs.’ (YPAG member)*

And some comment on the reporting of the interventions:

> *‘I think that what sounds interesting or boring when reading the research proposals could be very different if actually taking part in the studies.’ (YPAG member)*

## Discussion

Our extensive engagement with CYP, teachers and public health professionals led to the development of a novel coding scheme that we used to code 254 interventions in 210 randomized trials. Our consultations highlighted themes such as the recipient of the intervention (e.g. child, family, school, community); aspects of setting (e.g. home vs school vs community); duration and intensity of the intervention (e.g. low level intensity and long duration vs high level intensity and short duration); integration of the intervention (e.g. fully integrated in the curriculum vs intermediate vs not at all); choice and flexibility (e.g. children can choose the type of physical activity, whether the intervention can be implemented in a flexible manner); the ‘fun factor’ of the intervention (e.g. if the intervention is expected to be fun for everyone); resonance (e.g. the importance of role models or external professionals); mode of delivery of the intervention (e.g. by changing behaviour of the child vs educating the child vs changing the social and/or physical environment of the child).

A key strength was its iterative development through consultation with both recipients and implementers of obesity prevention interventions as well as with experts in the fields of obesity prevention and public health. Involvement of the project advisory group and its guidance in the design and implementation of the analytic framework was also highly beneficial. A particularly notable feature of our work was the involvement of CYP in both the development and the application of the analytic framework. They helped us determine the intervention characteristics included, and a group of 35 CYP performed the coding of all interventions in relation to the ‘fun factor’. Working with the CYP was mutually beneficial: both we and the CYP found the experience highly stimulating, and we believe the research was considerably improved by this partnership.

Challenges we encountered during the analytic framework development included overlap between some of the characteristics, finding appropriate wording of the questions and answers, and identification of characteristics that were unfeasible to code. Nonetheless, by iteratively applying changes to our various list of items to consider, we were able to refine the set of core features of interventions that we believe might have an impact on their effectiveness in preventing obesity in children. A limitation of the coding results is their dependency on the level of detail provided to describe the interventions. For most of the studies, the interventions were well-described and so we are confident that coding is accurate and reliable. However, some of the interventions were poorly described with limited information provided, an issue that likely affected the quality of the coding. Our feedback from the CYP involved in the coding also highlighted that some of the descriptions were not clear to them.

A limitation of this work relates to the demographic profile, particularly the socioeconomic status (SES), of the children and young people who took part in the workshops and ‘fun factor’ coding. Although we did not collect data on the SES of these children, our perception was that these children were most likely to come from middle-class families.

The approach described here should be suitable for application to other types of diverse and complex interventions and could be reproduced by other researchers (e.g. for evidence synthesis or intervention development). From the children involved in the coding, we learned the importance of ensuring that tasks offered to them are appropriately tailored to the age group. If conducting a similar exercise in the future, we would reserve additional resources for ensuring that intervention descriptions are edited to make them more understandable to the younger children.

In future work we will re-analyse the results of the randomized trials, feeding the results of our coding into a complex synthesis model. Through this analysis, using meta-regression-based methods within a Bayesian statistical framework, we will be able to evaluate the effect of each intervention component in producing a beneficial outcome in terms of prevention of obesity in children. The results of such analysis will potentially have an impact on the future development of interventions to prevent childhood obesity. Ultimately, the evidence produced by our main analysis may contribute to the reduction in childhood obesity.

## Supporting information

Apendix 6

## Data Availability

All data produced in the present work are contained in the manuscript

## Acknowledgements

We particularly thank the children, young people and parents participating in our workshops: Ameilia Holford, Amelie Low, Daniel Seretny, Elaine S Seretny, Francesca Quick, George Thomas, Havi Carel, Karen Low, Lucy Naser, Lucy Thomas, Nino Faber Gray, Riya Baghirathan Nicholls. We are also indebted to the children and young people who contributed to coding: Abdi Ali, Alice Matthews, Amaani Isse, Amelie Low, Amiira Isse, Archie Cazalet, Arwen Sofia Sequeira White Wandschneider, Ayaan Isse, Elizabeth Sheldrick, George Thomas, Harry Allbless Roberts, Hudda Mahamed, Isaac Hodge, Isaac Tregidgo, Jake Andrews, Joe Freer, Joseph Thomas, Katie Bond, Lottie Freer, Lowenna Negus, Matilda Hodge, Matilda Tregidgo, Max Andrews, Millie Freer, Oliver Leary, Oscar Lopez-Cottrell, Rahma Farah, Riya Baghirathan Nicholls, Shafia Mahamed, Sol Okasha, Sophie Phillips, Stanley Thomas, Tejas Ramanan, Theo Donnelly.

We thank the following teachers for participating in workshops: Stephen Breeze, Joanne Davies, Robert Davies, Gill Hampton, Martin Holmes, Emily Proffit, Gill Richards. We thank the following public health professionals participating in workshops and informal discussions: Justine Womack (OHID SW), Penny Marno (Swindon Borough Council), Lewis Bird (Swindon Borough Council), Sarah Amos (South Gloucestershire Council), Georgie MacArthur (North Somerset Council), Caoimhe Gowran (Bristol City Council).

Finally, we acknowledge the invaluable input from our project advisory group members: Jeremy Grimshaw (University of Ottawa), Tom Trikalinos (Brown University), Miranda Pallan (University of Birmingham), Graham Moore (Cardiff University), Lesley Stewart (University of York).

## Author contributions

- FS: Project organization; Scoping review; Analytic framework content; Coding of studies; Data analysis; Drafting manuscript.
- ALD: Analytic framework content; Coding of studies; Critical comments on manuscript.
- JCP: Analytic framework content; Coding of studies
- ET: Scoping review; Analytic framework content; Coding of studies Critical comments on manuscript.
- MC: Project organization; Analytic framework content (young person perspective)
- ES: Project organization; Analytic framework content (young person perspective)
- LC: Project organization; Patient/public involvement expertise; Critical comments on manuscript.
- THMM: Analytic framework content; Coding of studies; Critical comments on manuscript.
- DMC: Analytic framework content; Coding of studies; Critical comments on manuscript.
- FBG: Analytic framework content (behavioural change expertise); Critical comments on manuscript.
- SI: Analytic framework content; Critical comments on manuscript.
- JDN: Analytic framework content; Critical comments on manuscript.
- JS: Analytic framework content; Critical comments on manuscript.
- RC: Analytic framework content; Critical comments on manuscript.
- CDS: Conception; Obtaining funding; Analytic framework content (physical activity, nutrition and obesity perspectives); Critical comments on manuscript.
- JPTH: Conception; Obtaining funding; Supervision; Project organization; Analytic framework content (methodological perspective); Coding of studies; Drafting manuscript.

## Notes

### Competing Interest Statement

The authors have declared no competing interest.

