## Appendix 1 for "A novel analytic framework to investigate differential effects of interventions to prevent obesity in children and young people"

**Appendix 1.** Preliminary logic model

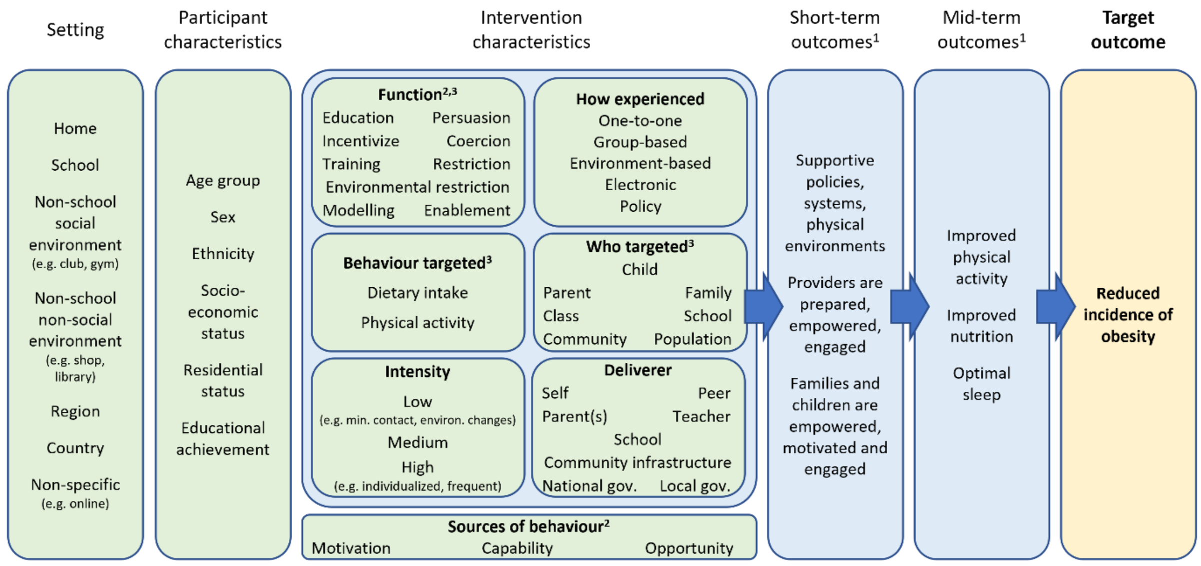


[1] Based on (O'Connor et al. 2015).
[2] Based on (Michie, van Stralen, and West 2011).
[3] More than one may apply.
