## Appendix 2 for "A novel analytic framework to investigate differential effects of interventions to prevent obesity in children and young people"

**Appendix 2**: Example of description of the interventions that we send to the children and young people for the coding of the 'fun factor'

### Strategies for prevention of obesity in children and young people: fun factor

**Batch: 1**

Please read the summary of the 10 strategies aiming to prevent children/young people from gaining excess weight and answer the questions in the survey for the strategies in the same order they appear below.

The survey can be found at https://sscm.onlinesurveys.ac.uk/strategies_fun_factor

| **Strategy 1** | |
|---|---|
| **Strategy ID** | **Adab 2018** |
| **Intended age group** | 6 |
| **Setting** | School + Home |
| **Strategy summary** | Several behaviour change strategies were employed to encourage increased physical activity and improved diet quality. School staff were provided with training and resources for intervention delivery. A termly family newsletter reinforced messages delivered through the various intervention components. The 12-month intervention encouraged healthy eating and physical activity, including a daily additional 30-minute school time physical activity opportunity, a six-week interactive skill-based programme in conjunction with Aston Villa football club, signposting of local family physical activity opportunities through mailouts every six months, and termly school led family workshops on healthy cooking skills. |

| **Strategy 2** | |
|---|---|
| **Strategy ID** | **Annesi 2016** |
| **Intended age group** | 7 |
| **Setting** | School (after school program) |
| **Strategy summary** | Youth Fit 4 Life use theory-based behavioural skills to support increased physical activity and healthy eating behaviours occurring both within and beyond after-school care time. It included highly structured daily session of 30 min/day of moderate-to vigorous physical activity and used cognitive-behavioral methods to encourage children to consume healthy foods and beverages.<br>The components of the daily sessions were similar and are indicated below:<br>• 5 min: active warm-up and focus upon a specific movement for the week (e.g., skipping) – 10 min: the day's assigned high-intensity activity^ (e.g., galloping tag);<br>• 10 min: alternate days of either a behavioral topic (e.g., positive self-talk^) or health topic (e.g., what is a grain?);<br>• 10 min: content reinforcement activity where the day's behavioral or health topic was bolstered by a structured physical activity (e.g., complete an assigned physical movement when a whole- vs. refined-grain food is named by a counsellor);<br>• 10 min: go-to game consisting of a moderate- to high-intensity game selected by the counsellor from an approved list. Posters supported the health topics, simple apparatus (e.g., cones, foam balls, hoops) supported the physical activities, and an activity sheet supported a participant-specific goal-setting process.<br>In an effort to obtain further support for the physical activity and nutrition behaviors, brief |

| | |
|---|---|
| | letters explaining what was recently emphasized within the program, and how it might be supported outside of school, were sent to parents/guardians weekly. In the fifth day of the week, the time allocated to physical activity was left to the discretion of the after-school care counsellor |

| **Strategy 3** | |
|---|---|
| **Strategy ID** | **Baranowski 2003** |
| **Intended age group** | 8 |
| **Setting** | Community + Telehealth + Web |
| **Strategy summary** | The intervention was a 4- week summer day camp, followed by an 8-week Internet-based program, plus one Saturday meeting for the girls. The intervention camp blended usual camp activities with activities specially designed for intervention. The specially designed interactive multimedia activities included buddy groups; camp cheers used as mnemonics for decision making, problem solving, and asking behaviours; training in dance; educational games targeted at increasing fruit juice and vegetable (FJV) intake and physical activity (PA); snack recipe preparation; and goal (called ‘‘challenges’’) setting and review. The weekly Website for the intervention girls included: a comic book with characters who attended the summer camp and faced and overcame hurdles in making lifestyle changes consistent with the dietary and PA goals; problem solving for challenges identified in the comic strips; review of attainment of previous week's goal; opportunities to set goals of 5 FJV servings/day, 5 glasses water/day, and 12,000 pedometer counts per day; a photo album of girls from the camp (both individual and group pictures); an ‘‘ask the expert’’ feature; and links to various Websites of interest to girls. Girls received weekly email and telephone reminders to log-on. The weekly Website for treatment parents included: a comic book in which a parent character commented on each frame of the child's comic. |

| **Strategy 4** | |
|---|---|
| **Strategy ID** | **Barnes 2015** |
| **Intended age group** | 9 |
| **Setting** | School (after school program) |
| **Strategy summary** | The MADE4Life program involved mothers and daughters attending weekly after-school 90-minute sessions over 8-weeks. The major focus of the mother-daughter physical activity (PA) sessions were fun active games, health-related fitness zumba, aerobics, pilates, yoga, rough and tumble play, and fundamental movement skills. Daughters' education sessions focused on developing an active lifestyle, benefits of PA and ways to reduce screen time. The 'daughter's booklet' contained weekly worksheets for daughters to complete with activities (eg, the importance of PA, fun ways to be active, reducing screen time). Daughters completed weekly 'pink slip' homework tasks that encouraged home PA with their mothers (eg, creating home-based fitness circuits). Pink slips were reviewed weekly by facilitators and daughters were rewarded with a 'scratch n smell' sticker to attach to a sticker chart. Mothers' education sessions consisted of evidence-based information on PA, behaviour change, role modeling and parenting strategies to support their daughter(s) PA. Sessions focused on the importance of mothers being a positive and active female role model. Mothers were given a 'mother's handbook' to file weekly session outlines and various resources that supported mother-daughter PA (eg, pedometers, skipping ropes). Mothers were encouraged to set SMART goals and self-monitor their daily PA using pedometers. |

| Strategy 5 | |
|---|---|
| **Strategy ID** | **Beech 2003** |
| **Intended age group** | 9 |
| **Setting** | School (after school program) |
| **Strategy summary** | The active interventions involved highly interactive weekly group sessions with either girls (child-targeted program) or parents/caregivers (parent-targeted program). Content focused on knowledge and behavior change skills to promote healthy eating and increased physical activity.<br>1. Child-targeted intervention ''GEMS Jamboree': girls participated in weekly, 90- minute intervention sessions for 12 weeks including ''Movin' It'' (physical activity component) and ''Munchin' It'' (nutrition component). Each weekly session concluded with a ''Taking It Home'' segment in which the concepts of the day were reviewed, incentives (small gifts) were given, and motivation for healthy eating and the maintenance of physical activity was provided.<br>2. Eating and Activity Skills for Youth (EASY) was conducted in a 12-week, 90-minute session format that included: a physical activity component of dancing (EASY Moves); a didactic nutrition segment (EASY Tips); and a segment alternating food preparation and nutrition-related games (EASY Fun). |

| Strategy 6 | |
|---|---|
| **Strategy ID** | **Bohnert 2013** |
| **Intended age group** | 9 |
| **Setting** | School (after school program) |
| **Strategy summary** | 30-week curriculum that includes 10 three-week modules. Each module covered a different sport, health, and leadership topic and was age-appropriate for early adolescents. Each session is led by trained coaches, is approximately 90 min in length, and is divided into two areas of focus: 50% covers physical instruction and energetic activity through traditional and non-traditional sports and fitness activities (e.g., rhythm and movement, soccer, flag football, volleyball, tennis, basketball, lacrosse, softball, golf, track and field) and 50% addresses age-appropriate health education, nutrition education, and leadership and life skills topics. The intervention focuses on enhancing girls' health literacy, empowering the girls to believe that they can make healthy choices as well as promoting self-control around health and life choices. A "girl of the day award" is given to the girl who worked the hardest at each session, along with a small prize. A healthy snack or meal is also provided at every session, along with take-home materials for families to reinforce program messages." |

| Strategy 7 | |
|---|---|
| **Strategy ID** | **Brandstetter 2012** |
| **Intended age group** | 8 |
| **Setting** | School (after school program) |
| **Strategy summary** | URMEL-ICE focused on health-promoting behaviour change in three areas: drinking sugar-sweetened beverages, spending time with screen media and being physically active. Main issues were the following: drinking water instead of soft drinks, discovering 'hidden' sugar in drinks, encouraging everyday physical activities, engaging in leisure activities without TV, learning about local sport and leisure facilities. The URMEL-ICE-intervention consists of material for 1 school year including 29 teaching units (each 30–60 min), 2 short blocks of physical activity exercise a day (each 5–7 min), 6 family homework lessons (tasks that |

| | |
|---|---|
| | cannot be accomplished by the child himself without the help of a parent) and materials for the training and information of the parents. |

| Strategy 8 | |
|---|---|
| **Strategy ID** | **Branscum 2013a** |
| **Intended age group** | 9 |
| **Setting** | School (after school program) |
| **Strategy summary** | During the 'Introduction & Purpose of lesson' the instructor introduced and reviewed the lesson's key objectives and covered necessary knowledge and skills in order to perform the behavior the lesson targeted. In the 'Benefits' module, children learned positive benefits associated with the health behavior being promoted and sketched a comic-panel showing such a benefit. Next, children participated in 'Role-Playing' with the instructor to practice skills learned in the lesson in two separate real-world examples: one with a parent or guardian, and one with a peer. Finally, during 'Goal Setting', the instructor reviewed the key objectives of the lesson, children have the opportunity to ask questions about the lesson, and children sketched comic-book panels of themselves setting goals, monitoring and self-rewarding themselves for engaging the behavior the lesson targeted. The behavioral objectives for each lesson of the experimental intervention were to enable children to: engage in no more than 2 hours of screen time per day (lesson 1), consume water and sugar-free drinks instead of sugar-sweetened beverages (lesson 2), participate in at least 60 minutes of physical activity per day (lesson 3), and consume 5 servings of fruits and vegetables per day. |

| Strategy 9 | |
|---|---|
| **Strategy ID** | **Branscum 2013b** |
| **Intended age group** | 9 |
| **Setting** | School (after school program) |
| **Strategy summary** | Each lesson consists of 4 modules: Introduction & Purpose of lesson, Comic-Book activity #1, Comic-Book activity #2 and Wrap-up. During the 'Introduction & Purpose of lesson' the instructor introduced and covered the lesson's key objectives and taught necessary knowledge and skills in order to perform the behavior the lesson targeted. In the 'Comic-Book activity #1' and 'Comic-Book activity #2' modules, children learned an aspect of comic-book creation and sequential art. Finally, during 'Wrap-up', the instructor reviewed the key objectives of the lesson, and children had the opportunity to ask questions about the lesson. The behavioral objectives for each lesson of the comparison intervention were to enable children to: engage in no more than 2 hours of screen time per day (lesson 1), consume water and sugar-free drinks instead of sugar-sweetened beverages (lesson 2), participate in at least 60 minutes of physical activity per day (lesson 3), and consume 5 servings of fruits and vegetables per day. |

| Strategy 10 | |
|---|---|
| **Strategy ID** | **Breheny 2020** |
| **Intended age group** | 9 |
| **Setting** | School |
| **Strategy summary** | The Daily Mile involves children doing an extra 15 min of activity by running or walking around a track within the school grounds. Schools map out a route or track in their school grounds. The intervention was carried out in lesson time at a time to suit each class during the school day, children left the classroom to run or walk around a predefined route within the school grounds for 15 min (on average equivalent to a distance of around 1 mile). The |

| | |
|---|---|
| | intervention was carried out in all but severe adverse weather conditions and required no change of clothing or footwear and was not a substitute for PE or break-times. Whilst advised as a daily activity, the frequency and duration were at the class teacher's discretion. Class teachers delivered the intervention and were permitted to adapt it for implementation, using motivational material such as certificates, or using it to facilitate learning within another subject area such as Maths. |
