## Appendix 3 for "A novel analytic framework to investigate differential effects of interventions to prevent obesity in children and young people"

**Appendix 3:** Refined logic model

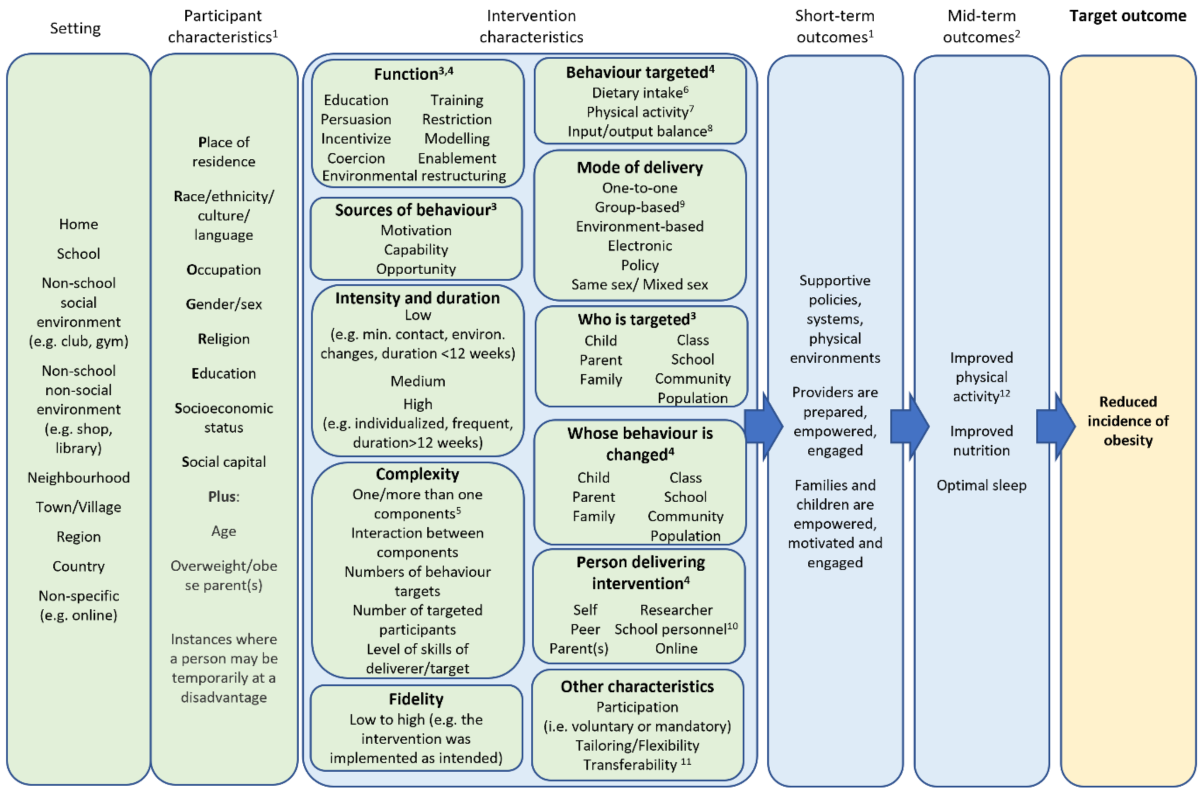


[1] Based on PROGRESS-Plus (O'Neill et al. 2014).
[2] Based on (O'Connor et al. 2015).
[3] Based on (Michie, van Stralen, and West 2011)
[4] More than one may apply.
[5] Integrated into a package and delivered as single or multiple intervention or delivered as bundle (Lewin et al. 2017).
[6] Including increase of healthy food and reduction of unhealthy food (O'Connor et al. 2015).
[7] Including increase of physical activity and reduction of sedentary behaviour (O'Connor et al. 2015).
[8] Metabolic processes modulators (e.g. sleep; stress).
[9] If family-based, are parents and/or siblings participants?
[10] Including teacher, nurse.
[11] The effect of intervention depends on context or setting in which it is implemented Lewin et al. 2017).
[12] Including reduction of sedentary behaviour.
