## Appendix 4 for "A novel analytic framework to investigate differential effects of interventions to prevent obesity in children and young people"

Appendix 4: Summary of themes addressed with children, young people and their parents

| Intervention characteristics | Contents of intervention |
| --- | --- |
| <ul><li>Fun</li><li>Casual</li><li>Enjoyable</li><li>Interesting</li><li>Interactive</li><li>Equitable – not excluding poorer families e.g. implementing local initiatives to get affordable family meals</li><li>Practical</li><li>Educational</li><li>Allowing for choice of activity</li><li>Integrated into existing 'systems'</li></ul>**Complexity**<ul><li>Simple (e.g. rap song as format that make it easy to remember the take-home message)</li><li>Easy to use (e.g. food boxes with instructions and weighed ingredients)</li></ul>**Mode of delivery**<ul><li>Group-based with class at school, friends, or as a family</li></ul>**Who targeted**<ul><li>Target parents/ family as well as the student</li><li>Targeting young children to instill habits from a young age – younger children more likely to do what they are told and have fewer stresses e.g. exams</li></ul> | ***Food-related***<ul><li>Teach children how to cook healthy food</li><li>Reduce the cost of healthy food in school and increase the cost of unhealthy food</li><li>Improve the quality of healthy food in school e.g. chopped fruit in pots, rather than gone off whole fruit</li><li>Raise awareness about nutrition and side effects of eating unhealthily</li><li>Limit how much unhealthy food can be bought in school</li><li>Reduce the amount of unhealthy food on offer in school</li><li>Increase availability of healthy snacks in the home</li><li>Involve children in preparing meals</li></ul>***Movement-related***<ul><li>Allow for choice of activity to let people do something they enjoy</li><li>Encourage people to do clubs outside of school</li><li>Increase physical activity after school</li><li>GP referral for exercise</li><li>Make exercise part of routine rather than something extra</li><li>Change uniform rules to reduce time wasted getting changed for PE and this as a barrier to enjoying PE in school</li></ul>***Setting***<ul><li>Impact the wider environment e.g. reducing canteen queues, improving healthy food offering in shops/ pharmacies, government</li></ul> |

**Person delivering intervention**

- Led by a credible teacher e.g. physical education (PE) teacher for exercise
- Role models – parents and famous people can influence what children do by the way they act
- Peer mentor/ ‘champion’

**Functions from logic model/ behaviour change wheel that came up in conversation**

- Restriction (e.g. reducing unhealthy food in school or limiting amount people can buy)
- Education
- Incentivization
- Coercion (briefly mentioned – e.g. to ensure people know the side effects of eating unhealthily e.g. on the NHS)
- Training
- Enablement
- Modelling (e.g. parents and famous people as role models)
- Environmental restructuring (e.g. changing types of food on offer in school, pharmacies, and increasing after school clubs)

initiatives, responsibility of food producers, places for children to socialize that aren’t fast food chains.

- Role models – parents, other children (more popular) and famous people can influence what children do by the way they act

***General/ other***

- Integrate into existing systems
- Incentives/ reward scheme
- Peer influence/ mentoring system
- Clearly demonstrate what the young person will get in return
- Use of apps
- Use a differentiated approach for young children vs older children
- Focus on mental health, meditation, self-awareness
- Improve school curriculum – more opportunities to explore more activities and do more cooking
- Consider the language used and the impact this has in different cultures – e.g. for an older generation in south Asian culture being ‘plump’ is positive. Saying that what is put in lunch boxes helps a child focus or do better at school may be more impactful than saying preventing obesity.
