## Appendix 5 for "A novel analytic framework to investigate differential effects of interventions to prevent obesity in children and young people"

**Appendix 5:** Analytic framework and coding manual

**Updated 8 March 2023**

| Item | Explanation | Question | Answer |
|---|---|---|---|
| **Setting** | This characteristic is a measure of the setting where the intervention is delivered in the sense of school vs home vs community.<br><br>School setting included after school programs based at school. Examples of community setting: club, gym, shop, library, health care centres.<br><br>Note that if the intervention is conducted within school facilities but set in the community we will answer **No** to school and **Yes** to community (e.g., an intervention in which families attend lessons and cooking classes, that uses the local school facilities (that is not necessarily the school that the participating children attend to).<br><br>It is possible to answer **Yes** to more than one of these questions. For example: an intervention that includes a school class on how to prepare healthy meals at home will be coded as **Yes** for school. If the intervention also includes delivery of a food box at home, we will also answer **Yes** to home.<br><br>General information for parents (e.g., flyer or newsletter) received at home as part of a wider strategy set in school or community is **No** to home.<br><br>An intervention that involves a significant component where the parent *receives instructions at home* to engage the child in behavioural changes (e.g. changes to meals or physical activities) is **Yes** to home. NB: If the instructions are delivered from the school (e.g. via the child) then this is **Yes** to school, **No** to home but **Yes** to 'home activity'. | Is the intervention delivered in a school (in full or in part)? | Yes/No |
| | | Is the intervention delivered in the home (in full or in part)? | Yes/No |
| | | Is the intervention delivered in the community or other non-school and non-home setting (in full or in part)? | Yes/No |
| | | Does the intervention include a home activity for the child? | Yes/No |

| | | | |
|---|---|---|---|
| | Examples of child **home activity**: homework (assigned according to the intervention protocol); cooking or games activities with parents.<br><br>If an intervention is entirely electronic and the study *does not specify* where the children should engage with the electronic activity (e.g. 'children must log in to a website at school') then answer **No** to all. Otherwise answer **Yes** to the specified location. | | |
| **Mode of delivery to the child** | This characteristic is a measure of how the child experiences the intervention. Although interventions may be delivered at various levels, the child will experience them in different ways, e.g. as an individualized intervention (e.g. a leaflet about healthy meals given to each student at school; a visit to an healthcare centre; homework with parents), through a group of children (e.g. school class or scout troop meeting), or otherwise.<br><br>Note: if the child experiences the intervention with the parents, we will code it as individual. An electronic intervention is coded as **Exclusively or mainly individually**.<br><br>If the intervention is delivered exclusively through electronic media (e.g. an app for exercising to use in the free time; a website to view at home) we will answer **Yes exclusively** to the second question. | How is the intervention delivered to the child? | Exclusively or mainly individually/ Both individually and as a group / Exclusively or mainly as a group |
| | | Is the intervention delivered to the child electronically? | Yes exclusively / Yes significantly / Yes as a minor component /No |
| **Realm targeted** | This characteristic is a measure of whether intervention seeks to change diet, activity (including increase in physical activity or decrease in sedentariness) or both. | Does the intervention aim to change diet? | Yes exclusively or substantially/Yes minimally/No |

| | | | |
|---|---|---|---|
| | Examples of changes in diet include introduction or replacement of food beverages with healthier options; re-organization of food display in the school canteen or in shops; education on healthy diet; cooking classes; healthy meal box for the family.<br><br>Examples of changes in activity includes intervention that increase physical activity (e.g. modified or additional physical activity classes at school) and interventions that reduce sedentary time at home (e.g. active video games).<br><br>We will answer **Yes exclusively/substantially** if the dietary or activity is the only realm targeted or if it is substantial in case of both dietary and activity interventions.<br><br>We will answer **Yes minimally** if the intervention is mainly one realm and there is a small component of the other realm (e.g. extension of the number of PA classes per week + a poster or leaflet about diet). | Does the intervention aim to change activity levels? | Yes exclusively or substantially/<br><br>Yes minimally/No |
| **Multifactorness / Dimensionality** | This characteristic is a measure of how non-simple/complex the intervention is. This includes how many ways the children are targeted, e.g., at multiple levels or in multiple phases.<br><br>Interventions targeting the children at multiple levels are those that use different strategies at the same time. Examples of multiple strategies interventions are intervention that include school lectures, school workshops, leaflets and homework.<br><br>Interventions targeting the children in multiple phases are interventions that use different strategies or settings at different time. A multi-phase intervention can also be an intervention with a more active phase followed by a less active “maintenance” phase or a “top-up” phase.<br><br>Interventions applied for a continuous period are interventions without breaks between the beginning and the end of the intervention (although school holidays don’t | Does the intervention use multiple strategies (three or more)?<br><br>Does the intervention applied have a single phase? | Yes/No<br><br>Yes/No |

| | | | |
|---|---|---|---|
| | count as a break in continuity of school-based interventions). Interventions applied for a discontinuous period are these with a break during the intervention (e.g. lectures delivered for 12 weeks/year for two years).<br><br>Examples of multiple strategy interventions delivered in multiple phases are interventions that include an initial series of school lectures at the end of which participants receive leaflets (phase 1) followed by a series of school workshops and homework (phase 2). | Is the intervention applied continuously? | Yes/No |
| **Peak intensity and duration** | This characteristic is a measure of how intensely the intervention is experienced by the child. Ideally this would cover the duration and frequency of the intervention.<br><br>In the case of a multiphase intervention, we will add the duration of similarly intense periods.<br><br>The answer to the question "**How many weeks does the intervention last?"** will be the number of weeks of active intervention. For example, an intervention delivered for 12 weeks/year over two school years will be coded as 24 weeks.<br><br>The answer to the question **"During how many weeks does the period of peak engagement with the intervention last?"** we will consider the duration of the period of high engagement, if there is a clear distinction between a period of high engagement and a period of low engagement (e.g., an active period and a maintenance period). | How many weeks does the intervention last (the period from baseline to end of intervention)? | Strictly numeric<br><br>*We are calculating the duration in weeks based on 4.33 weeks/month. In case of range duration (e.g. 16 to 20 weeks, we will take the mean=18)* |

| | | | |
|---|---|---|---|
| | Often the total duration and peak engagement period will be the same (unless phases of intensity are explicitly stated).<br><br>To answer the question "**What is the level of engagement with the children during the peak period?**" we will use the number of sessions of engagement per week as guidance:<br><br>• **High** engagement is typically one or more sessions of engagement with the children per week.<br>• **Low** engagement is typically less than one session of engagement with the children per week.<br><br>NB: These cut-offs are for guidance only. Sometimes the number of sessions per week will not be specified. Coders should use their judgement as to whether the intervention seems high or low intensity.<br><br>For permanent and transient environmental changes (e.g. changes in the display of food at the school canteen) this will be coded as **Low.** | During how many weeks does the period of peak engagement with the intervention last? | Strictly numeric<br><br>*See above* |
| | | What is the level of engagement with the children during the peak period? | High/Low |

| | | | |
|---|---|---|---|
| **Integration** | This characteristic is a measure of the extent to which the intervention is 'normalized' within the curriculum or normal habits. This measure provides an indication of how much 'extra effort' (by the provider and/or the recipient) would be required for the intervention to be successful.<br><br>Examples of **Yes** to intervention that is integrated: modification of physical activity classes; addition of, or replacement of regular school meals with, healthier options.<br><br>Examples of **Partially** answer is an intervention with a combination of integrated activities and something extra (e.g. after school program or homework).<br><br>Examples of **No** for an intervention that is not integrated at all): when the school needs to add something to an existing programme (e.g. an extra physical activity class extending school hours) or when the child needs to sign up for/agree to after-school classes.<br><br>After school programs (ASP): in case of ASP, the intervention is integrated if it seeks to change the content of an existing ASP and we will answer **Yes**; otherwise, it is not integrated, and we will answer **No**.<br><br>Electronic intervention: logging on to website is not integrated, receiving (and replying) to texts/messages/links is integrated. | Is the intervention integrated into the normal curriculum/ habits? | Yes/Partially (P)/No |
| **Flexibility** | This characteristic is a measure of the extent to which the intervention can be implemented flexibly, within the intervention protocol, e.g. an intervention is adapted to the particular classroom/household at teachers/parents' discretion.<br><br>Example of **Yes**: an intervention consisting in the replacement of regular meals with healthy meals where the healthy meals are decided by each participating school kitchen staff. Also, an intervention that is tailored to the specific characteristics of the participant (e.g. a dietary intervention that take into consideration what food the child likes or not). | Is the intervention designed to be implemented in a flexible manner/tailored to specific participants? | Yes/No |

| | | | |
|---|---|---|---|
| **Choice** | This characteristic is a measure of the extent to which participants (children) are free to make the intervention work for them.<br><br>Example of **Yes** is an intervention in which the child can choose which sport they do or which food to eat within the intervention.<br><br>Goal setting ? | Is choice for the child of activity/diet designed into the intervention? | Yes/Partially (P)/No |
| **Fun factor** | This characteristic is a measure of the extent to which the intervention is designed with the intention to be fun and whether children in the intended age group would find this strategy fun.<br><br>Examples of intervention that may look fun: game, song, play.<br><br>Example of intervention that may not look fun to all children: sport activity, cooking with the parents.<br><br>Example of intervention that may not look fun at all: a classroom lectures, replacement of sugar sweetened drinks with water.<br><br>Examples of intervention that children aged 5-11 year (but not an adolescent) will find fun: a song about healthy eating. Similarly, a video game intervention designed for older children (12- 18 years old) may not be fun for a 5-year-old child. | How enticing would you find this strategy? | Boring / Worse than neutral / Neutral / Better than neutral / Fun |
| | | How enticing do you think children in the intended age group would find this strategy? | Boring / Worse than neutral / Neutral / Better than neutral / Fun |
| **Resonance** | This characteristic is a measure of the extent to which the effectiveness of the intervention may depend on the degree of respect that young people have for the programme/deliverer, or on the credibility of the person delivering the intervention.<br><br>An example of **Yes** answer is an intervention in which the children are encouraged to do PE with an external PE teacher or coach. Also, an intervention in which workshops on healthy nutrition are delivered by a dietician or a nurse.<br><br>Other examples of role model are professional athletes (e.g., footballer), influencers, peers or older student. | Is the intervention experienced by children via someone external or unusual? | Yes/ No |

| | | | |
|---|---|---|---|
| | An example of **No** answer is an intervention in which the children are encouraged to do PA by a form teacher or a parent/career.<br><br>It will be a **Yes** answer if the intervention is delivered primarily by schoolteachers and one session is delivered for example by a dietician or a professional PA coach. | | |
| **Mechanism of action and recipient** | This characteristic is a measure of who is the direct recipient of the intervention (e.g., child, the teacher(s), parent(s), the child's environment or others) and how does the intervention aim to achieve a change in the child's behaviour.<br><br>Note that for complex interventions we may answer **Yes** to more than one question.<br><br>An example of an intervention that has a component that requires the child to participate is a session of physical activity or a workshop on healthy nutrition in which the children are involved in cooking a meal.<br><br>An example of an intervention that has a component of education or information is a provision of literature or lessons to *educate children* about the benefits of healthy eating/physical activity.<br><br>An example of an intervention that has a component aiming to change the social environment of the child at school or home is an intervention in which teachers are instructed to encourage children to change their dietary or activity behaviours or parents are educated on healthy food.<br><br>Training the teachers to deliver the intervention will normally be answered as **No.**<br><br>Examples of interventions that have a component aiming to change the physical environment of the child (at school or home) include the placement of healthy foods in the school canteen, provision of exercise equipment at school or in the community; drawing running tracks in the playground; changing the school meal menu. | Does the intervention have an explicit component that requires the child to participate? | Yes/No |
| | | Does the intervention have an explicit component of education/information provision for the child? | Yes/No |
| | | Does the intervention have an explicit component aiming to change the social environment of the child (e.g., at school or home)? | Yes/No |
| | | Does the intervention have an explicit component aiming to change the physical environment of the child (e.g., at school or home)? | Yes/No |

| | | | |
|---|---|---|---|
| **Commercial interests** | This characteristic is a measure of whether commercial interests are involved in the intervention (e.g. industry involvement).<br><br>An example of **Yes** answer is an intervention within a study that was funded by industry (e.g. food industry, manufacturer of sport equipment), even if the authors stated there were no conflict of interests. | Are commercial interests involved in the intervention? | Yes / No |
