## Supplementary material for "A novel analytic framework to investigate differential effects of interventions to prevent obesity in children and young people": Apendix 6

**Appendix 6**. Results of coding active interventions arms in the age 5-11 years and 12-18 years groups.

| Characteristic | Answer | Age 5-11 years (n = 188)* n (%)** | Age 12-18 years (n = 67)* n (%)** |
|---|---|---|---|
| **Setting** | | | |
| Delivered in school | Yes | 136 (72.3) | 44 (65.7) |
| | No | 52 (27.7) | 23 (34.3) |
| Delivered in the home | Yes | 38 (20.2) | 9 (13.4) |
| | No | 150 (79.8) | 58 (86.6) |
| Delivered in the community or other setting | Yes | 50 (26.6) | 22 (32.8) |
| | No | 138 (73.4) | 45 (67.2) |
| Includes a home activity | Yes | 71 (37.8) | 20 (29.9) |
| | No | 117 (62.2) | 47 (70.1) |
| **Mode of delivery** | | | |
| Delivered to the child | Individually | 26 (13.8) | 18 (26.9) |
| | Individually and as a group | 57 (30.3) | 21 (31.3) |
| | As a group | 105 (55.9) | 28 (41.8) |
| **Web component** | | | |
| Delivered electronically | Exclusively | 8 (4.3) | 8 (11.9) |
| | Significantly | 9 (4.8) | 8 (11.9) |
| | As a minor component | 15 (7.9) | 6 (9.0) |
| | No | 156 (83) | 45 (67.2) |
| **Realm targeted** | | | |
| Aims to change diet | Exclusively or substantially | 136 (72.3) | 50 (74.6) |
| | Minimally | 11 (5.9) | 2 (3.0) |
| | No | 41 (21.8) | 15 (22.4) |
| Aims to change activity | Exclusively or substantially | 161 (85.6) | 46 (68.7) |
| | Minimally | 4 (2.2) | 3 (4.4) |
| | No | 23 (12.2) | 18 (26.9) |
| **Multifactor-ness and dimensionality** | | | |
| Uses multiple strategies | Yes | 124 (66) | 37 (55.2) |
| | No | 64 (34) | 40 (44.8) |
| Applied in a single phase | Yes | 156 (83) | 51 (76.1) |
| | No | 32 (17) | 16 (23.9) |
| Applied for a continued period | Yes | 181 (96.3) | 65 (97.0) |
| | No | 7 (3.7) | 2 (3.0) |
| **Peak intensity and duration** | | | |
| Total duration | Mean weeks (SD) | 50.8 (45) | 32.4 (29.5) |
| Peak period duration | Mean weeks (SD) | 44.1 (44.1) | 26.8 (27.7) |
| Level of engagement with the child | High | 112 (59.6) | 40 (59.7) |
| | Low | 76 (40.4) | 27 (40.3) |

| | | | |
|---|---|---|---|
| **Integration** | | | |
| Integrated into the normal curriculum/habits | Completely | 85 (45.2) | 36 (53.7) |
| | Partially | 45 (23.9) | 10 (15.0) |
| | No | 58 (30.9) | 21 (31.3) |
| **Flexibility** | | | |
| Implemented in a flexible/tailored manner | Yes | 64 (34) | 22 (32.8) |
| | No | 124 (66) | 45 (67.2) |
| **Choice** | | | |
| Designed to have choice of activity and/or diet | Yes | 44 (23.4) | 22 (32.8) |
| | No | 144 (76.6) | 45 (67.2) |
| **Fun factor** | | | |
| How enticing for children in the intended age group | Fun | 120 (63.8) | 34 (50.7) |
| | Boring | 45 (23.9) | 26 (38.8) |
| | Neutral | 23 (12.2) | 7 (10.5) |
| **Resonance** | | | |
| Experienced via someone external or unusual | Yes | 96 (51.1) | 38 (56.7) |
| | No | 92 (48.9) | 29 (43.3) |
| **Mechanism of action and recipient** | | | |
| Participation | Yes | 131 (69.7) | 39 (58.2) |
| | No | 57 (30.30 | 28 (41.8) |
| Provision of education/information | Yes | 135 (71.8) | 55 (82.1) |
| | No | 53 (28.2) | 12 (17.9) |
| Change in child social environment | Yes | 137 (72.9) | 38 (56.7) |
| | No | 51 (27.1) | 29 (43.3) |
| Change in child physical environment | Yes | 63 (33.5) | 16 (23.9) |
| | No | 125 (66.5) | 51 (76.1) |
| **Commercial interests** | | | |
| Commercial interests in the trial and/or intervention | Yes | 21 (11.2) | 6 (9.0) |
| | No | 167 (88.8) | 61 (91.0) |

*One intervention conducted in both age groups.

**Except for duration which is reported as mean and standard deviation (SD).
