## Appendix 7 for "A novel analytic framework to investigate differential effects of interventions to prevent obesity in children and young people"

**Appendix 6:** Interventions coding full data set

| General informations | | | | |
|---|---|---|---|---|
| **Study ID** | **Age group** | **Arm** | **Comparison** | **Is the intervention delivered in a school (in full or in part)?** ***Yes/No*** |
| **Adab 2018** | 5-11 | WAVES | control | Yes |
| **Amaro 2006** | 12-18 | Kaledo | control | Yes |
| **Andrade 2014** | 12-18 | ACTIVAL | control | Yes |
| **Annesi 2016** | 5-11 | Youth Fit 4 Life | control | Yes |
| **Annesi 2017** | 5-11 | Youth Fit 4 Life | control | Yes |
| **Arlinghaus 2021** | 12-18 | FLOW-PA | control | Yes |
| **Baranowski 2003** | 5-11 | GEMS-FFFP | control | No |
| **Baranowski 2011** | 5-11 | Video Games | active | No |
| **Baranowski 2011** | 5-11 | Control | active | No |
| **Barbeau 2007** | 5-11 | Intervention | control | Yes |
| **Barnes 2015** | 5-11 | MADE4Life | control | No |
| **Barnes 2021** | 5-11 | SWAP IT interventic | control | Yes |
| **Barnes 2021** | 5-11 | Physically Active Ch | control | Yes |
| **Barnes 2021** | 5-11 | SWAP IT + PACE Cor | control | Yes |
| **Bayne-Smith 2004** | 12-18 | PATH | control | Yes |
| **Beech 2003** | 5-11 | Child targeted | control | No |
| **Beech 2003** | 5-11 | Parent targeted | control | No |
| **Black 2010** | 12-18 | Challenge! | control | No |
| **Bogart 2016** | 12-18 | SNaX | control | Yes |
| **Bohnert 2013** | 5-11 | GIG | control | Yes |
| **Bonsergent 2013** | 12-18 | Education | control | Yes |
| **Bonsergent 2013** | 12-18 | Environment | control | Yes |
| **Bonsergent 2013** | 12-18 | Screening and care | control | Yes |
| **Brandstetter 2012** | 5-11 | Intervention | control | Yes |
| **Branscum 2013** | 5-11 | Comics for Health | active | Yes |
| **Branscum 2013** | 5-11 | Knowledge based ir | active | Yes |
| **Breheny 2020** | 5-11 | Daily Mile | control | Yes |
| **Brito Beck da Silva 2019** | 12-18 | StayingFit | control | Yes |
| **Brown 2013** | 5-11 | Intervention | control | No |
| **Caballero 2003** | 5-11 | Intervention | control | Yes |
| **Cao 2015** | 5-11 | Intervention | control | Yes |
| **Chai 2019** | 5-11 | Telehealth | control | No |
| **Chai 2019** | 5-11 | Telehealth + SMS | control | No |
| **Chen 2010** | 5-11 | Intervention | control | No |
| **Chen 2011** | 12-18 | Web ABC | active | No |
| **Chen 2011** | 12-18 | Attention control | active | No |

| | | | | |
|---|---|---|---|---|
| **Choo 2020** | 5-11 | The Three-Healthy I | control | No |
| **Clemes 2020** | 5-11 | Stand Out in Class | control | Yes |
| **Coleman 2012** | 5-11 | Healthy ONES | control | Yes |
| **Crespo 2012** | 5-11 | APN Fam-only | control | No |
| **Crespo 2012** | 5-11 | APN Comm-only | control | Yes |
| **Crespo 2012** | 5-11 | APN Fam + Comm | control | Yes |
| **Cunha 2013** | 5-11 | PAPPAS | control | Yes |
| **Damsgaard 2014** | 5-11 | NND | control | Yes |
| **Davis 2021** | 5-11 | TX Sprouts | control | Yes |
| **De Bock 2013** | 5-11 | Ene mene fit | control | Yes |
| **de Greeff 2016** | 5-11 | F&V | control | Yes |
| **De Heer 2011** | 5-11 | Intervention | control | Yes |
| **de Ruyter 2012** | 5-11 | Intervention | control | Yes |
| **Dewar 2013** | 12-18 | NEAT GIRLS | control | Yes |
| **Diaz-Castro 2021** | 5-11 | PA intervention | control | Yes |
| **Donnelly 2009** | 5-11 | PAAC | control | Yes |
| **Drummy 2016** | 5-11 | Intervention | control | Yes |
| **Duncan 2019** | 5-11 | Healthy Homework | control | Yes |
| **Dunker 2018** | 12-18 | Brazilian New Move | control | Yes |
| **Ebbeling 2006** | 12-18 | Beverage interventi | control | No |
| **El Ansari 2010** | 12-18 | PA Intervention | control | Yes |
| **Elder 2014** | 5-11 | MOVE/me Muevo | control | No |
| **Ezendam 2012** | 12-18 | FATaintPHAT | control | Yes |
| **Fairclough 2013** | 5-11 | CHANGE! | control | Yes |
| **Farmer 2017** | 5-11 | PLAY | control | Yes |
| **Ford 2013** | 5-11 | Intervention | control | Yes |
| **Foster 2008** | 5-11 | SNPI | control | Yes |
| **French 2011** | 12-18 | Take Action | control | No |
| **Fulkerson 2010** | 5-11 | HOME | control | No |
| **Fulkerson 2015** | 5-11 | Intervention | control | No |
| **Fulkerson 2022** | 5-11 | NU-Home | control | No |
| **Gentile 2009** | 5-11 | SWITCH | control | Yes |
| **Greve 2015** | 5-11 | Intervention | control | Yes |
| **Griffin 2019** | 5-11 | Intervention | control | No |
| **Grydeland 2014** | 5-11 | HEIA | control | Yes |
| **Gustafson 2019** | 12-18 | Go Big and Take It H | control | No |
| **Ha 2021** | 5-11 | Active 1 + FUN | control | Yes |
| **Habib-Mourad 2014** | 5-11 | Health-E-PALS | control | Yes |
| **Habib-Mourad 2020** | 5-11 | Intervention | control | Yes |
| **Haerens 2006** | 12-18 | I+P | control | Yes |
| **Haerens 2006** | 12-18 | I | control | Yes |
| **Haire-Joshu 2010** | 5-11 | PARADE | control | No |
| **Han 2006** | 5-11 | Intervention | control | Yes |
| **Hannon 2018** | 5-11 | Intervention 2 | active | No |
| **Hannon 2018** | 5-11 | Intervention 1 | active | No |
| **Harrington 2018** | 12-18 | Girls Active | control | Yes |
| **HEALTHY Study Group 2010** | 5-11 | HEALTHY | control | Yes |
| **Hendrie 2011** | 5-11 | Intervention | control | No |
| **Hendy 2011** | 5-11 | KCP | control | Yes |
| **Hollis 2016** | 12-18 | Physical Activity 4 E | control | Yes |

| | | | | |
|---|---|---|---|---|
| **Hopper 2005** | 5-11 | Intervention | control | Yes |
| **Hovell 2018** | 12-18 | PAN | control | No |
| **Howe 2011** | 5-11 | Intervention | control | Yes |
| **Hull 2018** | 5-11 | Intervention | control | No |
| **Ickovics 2019** | 5-11 | Nutrition only | control | Yes |
| **Ickovics 2019** | 5-11 | PA only | control | Yes |
| **Ickovics 2019** | 5-11 | Nutrition and PA | control | Yes |
| **Isensee 2018** | 12-18 | Lauft program | control | Yes |
| **Jago 2006** | 12-18 | 5-a-day achievemer | active | No |
| **Jago 2006** | 12-18 | Fit for life | active | No |
| **James 2004** | 5-11 | CHOMPS | control | Yes |
| **Jansen 2011** | 5-11 | Intervention | control | Yes |
| **Jones 2015** | 5-11 | Wollongong SPORT | active | Yes |
| **Jones 2015** | 5-11 | Healthy lifestyle edu | active | Yes |
| **Kain 2014** | 5-11 | Intervention | control | Yes |
| **Keller 2009** | 5-11 | Intervention | control | No |
| **Kennedy 2018** | 12-18 | Resistance Training | control | Yes |
| **Keshani 2016** | 5-11 | Intervention | control | Yes |
| **Ketelhut 2022** | 5-11 | ExerCube interventi | control | Yes |
| **Khan 2014** | 5-11 | FITKids | control | Yes |
| **Kipping 2008** | 5-11 | Active for life year 5 | control | Yes |
| **Kipping 2014** | 5-11 | Active for life year 5 | control | Yes |
| **Klesges 2010** | 5-11 | Obesity prevention | control | No |
| **Kobel 2017** | 5-11 | Intervention | control | Yes |
| **Kocken 2016** | 5-11 | Intervention | control | Yes |
| **Kovalskys 2016** | 5-11 | SALTEN! | control | Yes |
| **Kriemler 2010** | 5-11 | KISS | control | Yes |
| **Kubik 2021** | 5-11 | SNAPSHOT | control | No |
| **Kuhlemeier 2022** | 12-18 | ACTION-PAC | control | Yes |
| **Kuroko 2020** | 12-18 | COOK (Create Our C | control | No |
| **Lappe 2017** | 12-18 | Dairy diet | active | No |
| **Lappe 2017** | 12-18 | Control | active | No |
| **Lau 2016** | 5-11 | Active Video Game | control | Yes |
| **Lazaar 2007** | 5-11 | Intervention | control | Yes |
| **Leme 2018** | 12-18 | H3-G-Brazil group | control | Yes |
| **Lent 2014** | 5-11 | Healthy Corner Stor | control | Yes |
| **Levy 2012** | 5-11 | Intervention | control | Yes |
| **Li 2010** | 5-11 | Happy 10 program | control | Yes |
| **Li 2019** | 5-11 | CHIRPY DRAGON | control | Yes |
| **Lichtenstein 2011** | 5-11 | GiZu Prevention Pro | control | Yes |
| **Liu 2019** | 5-11 | Intervention | control | Yes |
| **Liu 2022** | 5-11 | DECIDE - Children | control | Yes |
| **Llargues 2012** | 5-11 | Intervention | control | Yes |
| **Lloyd 2018** | 5-11 | HeLP | control | Yes |
| **Lubans 2021** | 12-18 | B2L | control | Yes |
| **Luszczynska 2016b** | 12-18 | Planning | control | Yes |
| **Luszczynska 2016b** | 12-18 | Self-efficacy | control | Yes |
| **Magnusson 2012** | 5-11 | Intervention | control | Yes |
| **Marcus 2009** | 5-11 | STOPP | control | Yes |
| **Martinez-Vizcaino 2014** | 5-11 | MOVI-2 | control | Yes |

| | | | | |
|---|---|---|---|---|
| **Martinez-Vizcaino 2020** | 5-11 | MOVI-Kids | control | Yes |
| **Martinez-Vizcaino 2022** | 5-11 | MOVI-daFIT! | control | Yes |
| **Melnyk 2013** | 12-18 | COPE | control | Yes |
| **Meng 2013 (Beijing)** | 5-11 | Happy 10 | control | Yes |
| **Meng 2013 (Beijing)** | 5-11 | Nutrition education | control | Yes |
| **Mihas 2010** | 12-18 | VYRONAS | control | Yes |
| **Morgan 2011** | 5-11 | HDHK program | control | No |
| **Morgan 2014** | 5-11 | HDHK program | control | No |
| **Morgan 2019** | 5-11 | DADEE | control | No |
| **Muller 2016** | 5-11 | Leipzig School Proje | control | Yes |
| **Muller 2019** | 5-11 | DASH | control | Yes |
| **Muzaffar 2019** | 5-11 | PAWS Peer led | active | Yes |
| **Muzaffar 2019** | 5-11 | PAWS Adult led | active | Yes |
| **NCT00224887 2005** | 5-11 | FBC | active | No |
| **NCT00224887 2005** | 5-11 | Control | active | No |
| **NCT02067728 2014** | 5-11 | FNPA tool | control | No |
| **Nemet 2011a** | 5-11 | Intervention | control | Yes |
| **Nemet 2011b** | 5-11 | Intervention | control | Yes |
| **Neumark-Sztainer 2003** | 12-18 | New Move | control | Yes |
| **Neumark-Sztainer 2010** | 12-18 | New Moves | control | Yes |
| **Newton 2014** | 5-11 | Intensive interventi | active | No |
| **Newton 2014** | 5-11 | Minimal interventio | active | No |
| **Nicholl 2021** | 5-11 | Milky Way study | control | No |
| **Nollen 2014** | 5-11 | MT | active | No |
| **Nollen 2014** | 5-11 | Control | active | Yes |
| **Nyberg 2015** | 5-11 | Healthy School Star | control | Yes |
| **Nyberg 2016** | 5-11 | Healthy School Star | control | Yes |
| **O'Connor 2020** | 5-11 | PSNS | control | No |
| **Ooi 2021** | 12-18 | SwitchURsip | control | Yes |
| **Paineau 2008** | 5-11 | Group A | control | Yes |
| **Paineau 2008** | 5-11 | Group B | control | Yes |
| **Papadaki 2010** | 12-18 | LP/LGI | control | No |
| **Papadaki 2010** | 12-18 | LP/HGI | control | No |
| **Papadaki 2010** | 12-18 | HP/LGI | control | No |
| **Papadaki 2010** | 12-18 | HP/HGI | control | No |
| **Pate 2005** | 12-18 | LEAP | control | Yes |
| **Pena 2021** | 5-11 | Intervention | active | Yes |
| **Pena 2021** | 5-11 | Control | active | Yes |
| **Peralta 2009** | 12-18 | FILA | control | Yes |
| **Pfeiffer 2019** | 12-18 | Girls on the Move | control | Yes |
| **Prins 2012** | 12-18 | YouRAction | control | Yes |
| **Prins 2012** | 12-18 | YouRAction+e | control | Yes |
| **Puder 2011** | 5-11 | Intervention | control | Yes |
| **Ramirez-Rivera 2021** | 5-11 | Planet Nutrition | control | Yes |
| **Reesor 2019** | 12-18 | Weight managemer | control | Yes |
| **Rerksuppaphol 2017** | 5-11 | Control | active | Yes |
| **Rerksuppaphol 2017** | 5-11 | Internet-based prog | active | Yes |
| **Rhodes 2019** | 5-11 | Planning and educa | active | No |
| **Rhodes 2019** | 5-11 | Education | active | No |
| **Robinson 2003** | 5-11 | Intervention | active | No |

| | | | | |
|---|---|---|---|---|
| **Robinson 2003** | 5-11 | Control | active | No |
| **Robinson 2010** | 5-11 | Intervention | active | No |
| **Robinson 2010** | 5-11 | Control | active | No |
| **Rodearmel 2006** | 12-18 | Experimental | control | No |
| **Rosario 2012** | 5-11 | Intervention | control | Yes |
| **Rosenkranz 2010** | 5-11 | SNAP | control | No |
| **Rush 2012** | 5-11 | Project Energize | control | Yes |
| **Sacchetti 2013** | 5-11 | Intervention | control | Yes |
| **Safdie 2013** | 5-11 | Basic | control | Yes |
| **Safdie 2013** | 5-11 | Basic plus | control | Yes |
| **Sahota 2001** | 5-11 | APPLES program | control | Yes |
| **Sahota 2019** | 5-11 | Intervention | control | Yes |
| **Salmon 2022** | 5-11 | PA-I | control | Yes |
| **Salmon 2022** | 5-11 | SB-I | control | Yes |
| **Salmon 2022** | 5-11 | PA-I + SB-I | control | Yes |
| **Santos 2014** | 5-11 | Healthy Buddies | control | Yes |
| **Schreier 2013** | 12-18 | Volunteering interv | control | No |
| **Seguin-Fawler 2021** | 5-11 | CO-CSA + Education | control | No |
| **Sekhavat 2014** | 5-11 | Intervention | control | No |
| **Sgambato 2019** | 5-11 | Intervention | control | Yes |
| **Sherwood 2019** | 5-11 | Intervention | control | No |
| **Shin 2015** | 12-18 | The Baltimore Healt | control | No |
| **Shomaker 2019** | 12-18 | Mindfullness group | control | No |
| **Sichieri 2008** | 5-11 | Intervention | control | Yes |
| **Siegrist 2013** | 5-11 | JuvenTUM | control | Yes |
| **Siegrist 2018** | 5-11 | Intervention | control | Yes |
| **Simon 2008** | 5-11 | ICAP | control | Yes |
| **Simons 2015** | 12-18 | Active video game i | control | No |
| **Singh 2009** | 12-18 | DOiT | control | Yes |
| **Smith 2014** | 12-18 | ATLAS | control | Yes |
| **Spiegel 2006** | 5-11 | WAY | control | Yes |
| **Stettler 2015** | 5-11 | Beverage-only | control | No |
| **Stettler 2015** | 5-11 | Multiple behaviors | control | No |
| **Stolley 1997** | 5-11 | Intervention | control | No |
| **Story 2003** | 5-11 | Girlfriends for KEEP | control | Yes |
| **Story 2012** | 5-11 | Bright Start | control | Yes |
| **Takacs 2020** | 12-18 | Nutrition interventi | control | Yes |
| **Tanskey 2017** | 5-11 | 100 Miles club | control | Yes |
| **Tanskey 2017** | 5-11 | Just Move | control | Yes |
| **Telford 2012** | 5-11 | Specialist-taught PE | control | Yes |
| **Tessier 2008** | 5-11 | REGU'LAPS | active | Yes |
| **Tessier 2008** | 5-11 | Active Control | active | Yes |
| **Thivel 2011** | 5-11 | Intervention | control | Yes |
| **Topham 2021** | 5-11 | FL | control | No |
| **Topham 2021** | 5-11 | FL + FD | control | No |
| **Topham 2021** | 5-11 | FL + PG | control | Yes |
| **Topham 2021** | 5-11 | FL + FD + PG | control | Yes |
| **van de Berg 2020** | 5-11 | WAT+LGEG | control | Yes |
| **van de Berg 2020** | 5-11 | LGEG | control | Yes |
| **van de Berg 2020** | 5-11 | WAT | control | Yes |

| | | | | |
|---|---|---|---|---|
| **Velez 2010** | 12-18 | Resistance training | control | Yes |
| **Viggiano 2015** | 12-18 | Kaledo | control | Yes |
| **Viggiano 2018** | 5-11 | Intervention | control | Yes |
| **Vizcaino 2008** | 5-11 | MOVI | control | Yes |
| **Wang 2012** | 5-11 | Intervention | control | Yes |
| **Wang 2018** | 5-11 | HLP-YOG interventi | control | Yes |
| **Weeks 2012** | 12-18 | POWER PE-Total IT | control | Yes |
| **Wendel 2016** | 5-11 | Intervention | control | Yes |
| **White 2019** | 5-11 | Intervention | control | No |
| **Whittemore 2013** | 12-18 | HEALTH[e]TEEN+CS | active | Yes |
| **Whittemore 2013** | 12-18 | HEALTH[e]TEEN | active | Yes |
| **Wieland 2018** | 12-18 | The Healthy Immigr | control | No |
| **Wilksch 2015** | 12-18 | Life Smart | control | Yes |
| **Williamson 2012** | 5-11 | PP | control | Yes |
| **Williamson 2012** | 5-11 | PP+PS | control | Yes |
| **Xu 2015** | 5-11 | CLICK programme | control | Yes |
| **Xu 2017 (5 other cities)** | 5-11 | Comprehensive | control | Yes |
| **Yin 2012** | 5-11 | Fitkid Project | control | Yes |

dren and young people

| Setting | | |
|---|---|---|
| **Is the intervention delivered in the home (in full or in part)?** ***Yes/No*** | **Is the intervention delivered in the community or other non-school and non-home setting (in full or in part)?** ***Yes/No*** | **Does the intervention include a home activity?** ***Yes/No*** |
| No | Yes | Yes |
| No | No | No |
| No | No | No |
| No | No | No |
| No | No | No |
| No | No | No |
| Yes | Yes | Yes |
| Yes | No | Yes |
| Yes | No | Yes |
| No | No | No |
| No | Yes | Yes |
| Yes | No | No |
| No | No | No |
| Yes | No | No |
| No | No | Yes |
| No | Yes | No |
| No | Yes | No |
| Yes | Yes | Yes |
| No | No | Yes |
| Yes | No | Yes |
| No | No | No |
| No | No | No |
| No | Yes | No |
| No | No | Yes |
| No | No | No |
| No | No | No |
| No | No | No |
| No | No | No |
| No | Yes | Yes |
| No | No | Yes |
| Yes | No | Yes |
| No | No | No |
| No | No | No |
| Yes | Yes | Yes |
| Yes | Yes | Yes |
| Yes | Yes | Yes |

| | | |
|---|---|---|
| Yes | Yes | No |
| No | No | No |
| No | No | No |
| Yes | No | No |
| No | Yes | Yes |
| Yes | Yes | Yes |
| No | No | Yes |
| No | No | No |
| No | No | No |
| No | Yes | No |
| No | No | No |
| No | No | No |
| No | No | Yes |
| No | No | Yes |
| No | No | No |
| No | No | No |
| No | No | No |
| No | No | Yes |
| No | No | No |
| Yes | No | No |
| No | No | No |
| Yes | Yes | Yes |
| No | No | No |
| No | No | Yes |
| No | No | No |
| No | No | No |
| No | No | No |
| Yes | Yes | Yes |
| No | Yes | No |
| Yes | Yes | Yes |
| Yes | Yes | Yes |
| Yes | Yes | Yes |
| No | No | Yes |
| No | Yes | Yes |
| No | No | No |
| No | No | No |
| No | No | No |
| No | No | No |
| No | No | No |
| No | No | Yes |
| No | No | No |
| No | Yes | Yes |
| No | No | No |
| Yes | Yes | Yes |
| No | Yes | Yes |
| No | No | No |
| No | No | No |
| Yes | No | No |
| No | No | Yes |
| No | No | No |

| | | |
|---|---|---|
| No | No | Yes |
| No | Yes | No |
| No | No | No |
| Yes | Yes | Yes |
| No | No | No |
| No | No | No |
| No | No | No |
| No | No | No |
| No | Yes | Yes |
| No | Yes | Yes |
| No | No | No |
| No | No | Yes |
| No | No | Yes |
| No | No | No |
| No | No | No |
| No | Yes | No |
| No | No | No |
| No | No | No |
| No | No | No |
| No | No | No |
| No | No | No |
| No | No | Yes |
| No | Yes | No |
| No | No | Yes |
| Yes | No | Yes |
| No | No | No |
| No | No | Yes |
| Yes | Yes | Yes |
| No | No | No |
| Yes | Yes | Yes |
| No | Yes | No |
| No | Yes | No |
| No | No | No |
| No | No | No |
| No | No | No |
| No | Yes | No |
| No | No | No |
| No | No | No |
| No | No | Yes |
| No | No | No |
| No | No | Yes |
| No | No | Yes |
| No | No | No |
| No | No | Yes |
| No | No | Yes |
| No | No | No |
| No | No | No |
| No | No | No |
| No | No | No |
| No | No | No |

| | | |
|---|---|---|
| Yes | No | No |
| No | No | No |
| No | No | Yes |
| No | No | No |
| No | No | No |
| No | No | No |
| No | Yes | Yes |
| No | Yes | Yes |
| No | Yes | Yes |
| No | No | No |
| No | No | No |
| No | No | No |
| No | No | No |
| Yes | No | No |
| No | Yes | No |
| No | Yes | No |
| No | No | Yes |
| No | No | Yes |
| No | No | No |
| No | No | No |
| Yes | No | No |
| Yes | No | No |
| Yes | No | No |
| No | No | No |
| No | No | No |
| No | No | Yes |
| No | No | Yes |
| No | Yes | Yes |
| No | No | No |
| Yes | Yes | No |
| Yes | Yes | No |
| No | Yes | No |
| No | Yes | No |
| No | Yes | No |
| No | Yes | No |
| No | No | No |
| No | No | Yes |
| No | No | Yes |
| No | No | No |
| No | No | No |
| No | No | Yes |
| No | Yes | Yes |
| No | No | Yes |
| No | No | No |
| No | No | No |
| No | No | No |
| No | No | No |
| Yes | No | No |
| Yes | No | No |
| Yes | Yes | Yes |

| | | |
|---|---|---|
| No | Yes | No |
| Yes | Yes | Yes |
| No | Yes | No |
| No | Yes | Yes |
| No | No | No |
| No | Yes | Yes |
| No | No | No |
| No | No | No |
| No | No | No |
| No | No | No |
| No | No | No |
| Yes | No | Yes |
| No | No | Yes |
| No | No | Yes |
| No | No | Yes |
| No | No | No |
| No | Yes | No |
| Yes | Yes | Yes |
| No | Yes | No |
| Yes | No | Yes |
| Yes | Yes | No |
| Yes | Yes | Yes |
| No | Yes | Yes |
| No | No | No |
| No | No | Yes |
| No | No | Yes |
| No | Yes | No |
| Yes | No | Yes |
| No | No | No |
| No | No | No |
| No | No | Yes |
| No | Yes | No |
| No | Yes | No |
| No | Yes | No |
| No | No | Yes |
| No | No | No |
| No | No | Yes |
| No | No | No |
| No | No | No |
| No | No | No |
| No | No | No |
| No | No | No |
| No | No | No |
| No | Yes | No |
| No | Yes | No |
| No | Yes | No |
| No | Yes | No |
| Yes | No | Yes |
| Yes | No | Yes |
| Yes | No | Yes |

| | | |
|---|---|---|
| No | No | No |
| No | No | No |
| No | No | No |
| No | No | No |
| No | No | No |
| No | No | Yes |
| No | No | No |
| No | No | No |
| No | Yes | Yes |
| No | No | No |
| No | No | No |
| Yes | Yes | No |
| No | No | No |
| No | No | Yes |
| No | No | Yes |
| No | No | No |
| No | No | No |
| No | No | No |

| Mode of delivery to the child | | Realm t |
|---|---|---|
| **How is the intervention delivered?** ***Exclusively or mainly individually/ Both individually and as a group / Exclusively or mainly as a group*** | **Is the intervention delivered electronically?** ***Yes exclusively/ Yes significantly/Yes as a minor component/No*** | **Does the intervention aim to change diet?** ***Yes exclusively or substantially / Yes minimally / No*** |
| Exclusively or mainly as a group | No | Yes exclusively/substantially |
| Exclusively or mainly as a group | No | Yes exclusively/substantially |
| Exclusively or mainly as a group | No | Yes exclusively/substantially |
| Exclusively or mainly as a group | No | Yes exclusively/substantially |
| Exclusively or mainly as a group | No | Yes exclusively/substantially |
| Exclusively or mainly as a group | No | No |
| Both individually and as a group | Yes significantly | Yes exclusively/substantially |
| Exclusively or mainly individually | Yes exclusively | Yes exclusively/substantially |
| Exclusively or mainly individually | Yes exclusively | Yes exclusively/substantially |
| Exclusively or mainly as a group | No | Yes minimally |
| Both individually and as a group | No | No |
| Both individually and as a group | No | Yes exclusively/substantially |
| Exclusively or mainly as a group | No | No |
| Both individually and as a group | No | Yes exclusively/substantially |
| Exclusively or mainly as a group | No | Yes minimally |
| Exclusively or mainly as a group | No | Yes exclusively/substantially |
| Exclusively or mainly as a group | No | Yes exclusively/substantially |
| Exclusively or mainly individually | No | Yes exclusively/substantially |
| Both individually and as a group | No | Yes exclusively/substantially |
| Both individually and as a group | No | Yes exclusively/substantially |
| Exclusively or mainly as a group | No | Yes exclusively/substantially |
| Exclusively or mainly as a group | No | Yes exclusively/substantially |
| Exclusively or mainly as a group | No | Yes exclusively/substantially |
| Exclusively or mainly individually | No | Yes exclusively/substantially |
| Exclusively or mainly as a group | No | Yes exclusively/substantially |
| Exclusively or mainly as a group | No | Yes exclusively/substantially |
| Exclusively or mainly as a group | No | No |
| Exclusively or mainly individually | Yes exclusively | Yes exclusively/substantially |
| Exclusively or mainly as a group | No | Yes exclusively/substantially |
| Exclusively or mainly as a group | No | Yes exclusively/substantially |
| Both individually and as a group | No | Yes exclusively/substantially |
| Exclusively or mainly individually | Yes exclusively | Yes exclusively/substantially |
| Exclusively or mainly individually | Yes significantly | Yes exclusively/substantially |
| Both individually and as a group | Yes as a minor component | Yes exclusively/substantially |
| Exclusively or mainly individually | Yes exclusively | Yes exclusively/substantially |
| Exclusively or mainly individually | Yes exclusively | Yes exclusively/substantially |

| | | |
|---|---|---|
| Exclusively or mainly as a group | No | Yes exclusively/substantially |
| Exclusively or mainly as a group | No | No |
| Both individually and as a group | No | Yes exclusively/substantially |
| Exclusively or mainly individually | No | Yes exclusively/substantially |
| Exclusively or mainly as a group | No | Yes exclusively/substantially |
| Both individually and as a group | No | Yes exclusively/substantially |
| Exclusively or mainly as a group | No | Yes exclusively/substantially |
| Exclusively or mainly as a group | No | Yes exclusively/substantially |
| Exclusively or mainly as a group | No | Yes exclusively/substantially |
| Exclusively or mainly as a group | Yes as a minor component | Yes minimally |
| Exclusively or mainly as a group | No | No |
| Exclusively or mainly as a group | No | Yes exclusively/substantially |
| Exclusively or mainly as a group | No | Yes exclusively/substantially |
| Both individually and as a group | Yes as a minor component | Yes exclusively/substantially |
| Exclusively or mainly as a group | No | No |
| Exclusively or mainly as a group | No | No |
| Exclusively or mainly as a group | No | No |
| Both individually and as a group | Yes as a minor component | Yes exclusively/substantially |
| Both individually and as a group | No | Yes exclusively/substantially |
| Exclusively or mainly individually | No | Yes exclusively/substantially |
| Exclusively or mainly as a group | No | No |
| Both individually and as a group | No | Yes exclusively/substantially |
| Exclusively or mainly individually | Yes exclusively | Yes exclusively/substantially |
| Exclusively or mainly as a group | No | Yes exclusively/substantially |
| Exclusively or mainly as a group | No | No |
| Exclusively or mainly as a group | No | No |
| Exclusively or mainly as a group | No | Yes exclusively/substantially |
| Both individually and as a group | No | Yes exclusively/substantially |
| Exclusively or mainly as a group | No | Yes exclusively/substantially |
| Both individually and as a group | No | Yes exclusively/substantially |
| Both individually and as a group | No | Yes exclusively/substantially |
| Both individually and as a group | No | Yes exclusively/substantially |
| Both individually and as a group | Yes significantly | Yes exclusively/substantially |
| Both individually and as a group | No | Yes exclusively/substantially |
| Both individually and as a group | Yes as a minor component | Yes exclusively/substantially |
| Exclusively or mainly individually | Yes exclusively | Yes exclusively/substantially |
| Exclusively or mainly as a group | Yes as a minor component | No |
| Exclusively or mainly as a group | No | Yes exclusively/substantially |
| Exclusively or mainly as a group | No | Yes exclusively/substantially |
| Both individually and as a group | Yes significantly | Yes exclusively/substantially |
| Both individually and as a group | Yes significantly | Yes exclusively/substantially |
| Exclusively or mainly individually | Yes significantly | Yes exclusively/substantially |
| Exclusively or mainly as a group | No | Yes exclusively/substantially |
| Both individually and as a group | No | Yes exclusively/substantially |
| Both individually and as a group | No | Yes exclusively/substantially |
| Exclusively or mainly as a group | No | No |
| Exclusively or mainly as a group | No | Yes exclusively/substantially |
| Exclusively or mainly individually | No | Yes exclusively/substantially |
| Both individually and as a group | No | Yes exclusively/substantially |
| Exclusively or mainly as a group | No | No |

| | | |
|---|---|---|
| Both individually and as a group | No | Yes exclusively/substantially |
| Exclusively or mainly individually | No | Yes exclusively/substantially |
| Exclusively or mainly as a group | No | Yes minimally |
| Both individually and as a group | No | Yes exclusively/substantially |
| Exclusively or mainly as a group | No | Yes exclusively/substantially |
| Exclusively or mainly as a group | No | Yes minimally |
| Exclusively or mainly as a group | No | Yes exclusively/substantially |
| Both individually and as a group | Yes significantly | No |
| Both individually and as a group | Yes significantly | Yes exclusively/substantially |
| Both individually and as a group | Yes significantly | No |
| Exclusively or mainly as a group | Yes as a minor component | Yes exclusively/substantially |
| Exclusively or mainly as a group | No | Yes exclusively/substantially |
| Both individually and as a group | No | No |
| Both individually and as a group | No | Yes exclusively/substantially |
| Exclusively or mainly as a group | No | Yes exclusively/substantially |
| Exclusively or mainly individually | No | Yes exclusively/substantially |
| Exclusively or mainly as a group | Yes significantly | No |
| Exclusively or mainly as a group | No | Yes exclusively/substantially |
| Exclusively or mainly individually | Yes exclusively | No |
| Exclusively or mainly as a group | No | Yes minimally |
| Exclusively or mainly as a group | No | Yes exclusively/substantially |
| Exclusively or mainly as a group | No | Yes exclusively/substantially |
| Exclusively or mainly as a group | No | Yes exclusively/substantially |
| Exclusively or mainly as a group | No | Yes exclusively/substantially |
| Both individually and as a group | No | Yes exclusively/substantially |
| Exclusively or mainly as a group | No | Yes minimally |
| Both individually and as a group | No | No |
| Both individually and as a group | No | Yes exclusively/substantially |
| Exclusively or mainly individually | No | Yes exclusively/substantially |
| Both individually and as a group | Yes as a minor component | Yes exclusively/substantially |
| Exclusively or mainly individually | No | Yes exclusively/substantially |
| Exclusively or mainly individually | No | Yes minimally |
| Exclusively or mainly as a group | Yes exclusively | No |
| Exclusively or mainly as a group | No | No |
| Both individually and as a group | Yes as a minor component | Yes exclusively/substantially |
| Both individually and as a group | Yes as a minor component | Yes exclusively/substantially |
| Exclusively or mainly as a group | No | Yes exclusively/substantially |
| Exclusively or mainly as a group | No | No |
| Both individually and as a group | No | Yes exclusively/substantially |
| Exclusively or mainly as a group | No | Yes exclusively/substantially |
| Exclusively or mainly as a group | Yes as a minor component | Yes exclusively/substantially |
| Both individually and as a group | Yes as a minor component | Yes exclusively/substantially |
| Exclusively or mainly as a group | No | Yes exclusively/substantially |
| Exclusively or mainly as a group | No | Yes exclusively/substantially |
| Exclusively or mainly as a group | Yes significantly | No |
| Both individually and as a group | No | Yes exclusively/substantially |
| Both individually and as a group | No | Yes exclusively/substantially |
| Exclusively or mainly as a group | No | Yes exclusively/substantially |
| Exclusively or mainly as a group | No | Yes exclusively/substantially |
| Exclusively or mainly as a group | No | No |

| | | |
|---|---|---|
| Exclusively or mainly as a group | Yes as a minor component | No |
| Exclusively or mainly as a group | No | No |
| Exclusively or mainly individually | No | Yes exclusively/substantially |
| Exclusively or mainly as a group | No | No |
| Exclusively or mainly as a group | No | Yes exclusively/substantially |
| Exclusively or mainly as a group | No | Yes exclusively/substantially |
| Exclusively or mainly as a group | No | Yes exclusively/substantially |
| Both individually and as a group | No | Yes exclusively/substantially |
| Both individually and as a group | No | No |
| Exclusively or mainly as a group | No | No |
| Exclusively or mainly as a group | No | No |
| Exclusively or mainly as a group | No | Yes exclusively/substantially |
| Exclusively or mainly as a group | No | Yes exclusively/substantially |
| Exclusively or mainly individually | Yes as a minor component | Yes exclusively/substantially |
| Exclusively or mainly as a group | No | Yes exclusively/substantially |
| Exclusively or mainly individually | No | Yes exclusively/substantially |
| Exclusively or mainly as a group | No | Yes exclusively/substantially |
| Exclusively or mainly as a group | No | Yes exclusively/substantially |
| Both individually and as a group | No | Yes exclusively/substantially |
| Both individually and as a group | No | Yes exclusively/substantially |
| Exclusively or mainly individually | No | No |
| Exclusively or mainly individually | No | Yes minimally |
| Exclusively or mainly individually | No | Yes exclusively/substantially |
| Exclusively or mainly individually | Yes exclusively | Yes exclusively/substantially |
| Exclusively or mainly individually | No | Yes exclusively/substantially |
| Both individually and as a group | No | Yes exclusively/substantially |
| Both individually and as a group | No | Yes exclusively/substantially |
| Exclusively or mainly as a group | No | Yes exclusively/substantially |
| Exclusively or mainly as a group | Yes as a minor component | Yes exclusively/substantially |
| Both individually and as a group | Yes significantly | Yes exclusively/substantially |
| Both individually and as a group | Yes significantly | Yes exclusively/substantially |
| Both individually and as a group | No | Yes exclusively/substantially |
| Both individually and as a group | No | Yes exclusively/substantially |
| Both individually and as a group | No | Yes exclusively/substantially |
| Both individually and as a group | No | Yes exclusively/substantially |
| Exclusively or mainly as a group | No | No |
| Both individually and as a group | Yes significantly | Yes exclusively/substantially |
| Exclusively or mainly individually | Yes exclusively | Yes exclusively/substantially |
| Exclusively or mainly as a group | No | Yes exclusively/substantially |
| Exclusively or mainly as a group | Yes as a minor component | No |
| Exclusively or mainly as a group | Yes exclusively | No |
| Exclusively or mainly as a group | Yes exclusively | No |
| Both individually and as a group | No | Yes exclusively/substantially |
| Exclusively or mainly as a group | No | Yes exclusively/substantially |
| Exclusively or mainly as a group | No | Yes exclusively/substantially |
| Exclusively or mainly individually | No | Yes exclusively/substantially |
| Exclusively or mainly individually | Yes exclusively | Yes exclusively/substantially |
| Exclusively or mainly individually | No | No |
| Exclusively or mainly individually | No | No |
| Both individually and as a group | No | Yes minimally |

| | | |
|---|---|---|
| Exclusively or mainly as a group | No | Yes exclusively/substantially |
| Both individually and as a group | No | Yes minimally |
| Exclusively or mainly as a group | No | Yes exclusively/substantially |
| Exclusively or mainly individually | No | Yes exclusively/substantially |
| Exclusively or mainly as a group | No | Yes exclusively/substantially |
| Both individually and as a group | No | Yes exclusively/substantially |
| Exclusively or mainly as a group | No | Yes exclusively/substantially |
| Exclusively or mainly as a group | No | No |
| Exclusively or mainly as a group | No | Yes exclusively/substantially |
| Exclusively or mainly as a group | No | Yes exclusively/substantially |
| Exclusively or mainly as a group | No | Yes exclusively/substantially |
| Both individually and as a group | Yes as a minor component | Yes exclusively/substantially |
| Both individually and as a group | No | No |
| Both individually and as a group | No | No |
| Both individually and as a group | No | No |
| Exclusively or mainly as a group | No | Yes exclusively/substantially |
| Exclusively or mainly individually | No | Yes exclusively/substantially |
| Both individually and as a group | Yes as a minor component | Yes exclusively/substantially |
| Exclusively or mainly individually | No | Yes exclusively/substantially |
| Both individually and as a group | Yes as a minor component | Yes exclusively/substantially |
| Exclusively or mainly individually | No | Yes exclusively/substantially |
| Exclusively or mainly individually | No | Yes exclusively/substantially |
| Exclusively or mainly as a group | No | Yes exclusively/substantially |
| Exclusively or mainly as a group | No | Yes exclusively/substantially |
| Both individually and as a group | No | Yes exclusively/substantially |
| Exclusively or mainly as a group | No | Yes exclusively/substantially |
| Exclusively or mainly as a group | No | No |
| Exclusively or mainly individually | Yes exclusively | No |
| Exclusively or mainly as a group | No | Yes exclusively/substantially |
| Both individually and as a group | Yes significantly | Yes exclusively/substantially |
| Both individually and as a group | Yes as a minor component | Yes exclusively/substantially |
| Exclusively or mainly individually | No | Yes exclusively/substantially |
| Exclusively or mainly individually | No | Yes exclusively/substantially |
| Exclusively or mainly as a group | No | Yes exclusively/substantially |
| Both individually and as a group | No | Yes exclusively/substantially |
| Exclusively or mainly as a group | No | Yes exclusively/substantially |
| Both individually and as a group | Yes as a minor component | Yes exclusively/substantially |
| Both individually and as a group | No | No |
| Exclusively or mainly as a group | No | No |
| Exclusively or mainly as a group | No | No |
| Exclusively or mainly as a group | No | No |
| Exclusively or mainly as a group | No | No |
| Exclusively or mainly as a group | No | No |
| Exclusively or mainly as a group | No | Yes exclusively/substantially |
| Exclusively or mainly as a group | No | Yes exclusively/substantially |
| Exclusively or mainly as a group | No | Yes exclusively/substantially |
| Exclusively or mainly as a group | No | Yes exclusively/substantially |
| Both individually and as a group | No | Yes exclusively/substantially |
| Both individually and as a group | No | Yes exclusively/substantially |
| Exclusively or mainly as a group | No | Yes minimally |

| | | |
|---|---|---|
| Exclusively or mainly as a group | No | No |
| Exclusively or mainly as a group | No | Yes exclusively/substantially |
| Exclusively or mainly as a group | No | Yes exclusively/substantially |
| Exclusively or mainly as a group | No | No |
| Exclusively or mainly as a group | No | Yes exclusively/substantially |
| Both individually and as a group | No | No |
| Exclusively or mainly as a group | No | No |
| Exclusively or mainly as a group | No | No |
| Both individually and as a group | Yes significantly | Yes exclusively/substantially |
| Exclusively or mainly as a group | No | Yes exclusively/substantially |
| Exclusively or mainly as a group | No | Yes exclusively/substantially |
| Exclusively or mainly individually | No | Yes exclusively/substantially |
| Exclusively or mainly as a group | No | Yes exclusively/substantially |
| Both individually and as a group | No | Yes exclusively/substantially |
| Both individually and as a group | Yes significantly | Yes exclusively/substantially |
| Exclusively or mainly as a group | No | Yes exclusively/substantially |
| Exclusively or mainly as a group | No | Yes exclusively/substantially |
| Exclusively or mainly as a group | No | Yes minimally |

| argeted | Multifactorness / Dimensionality | | |
|---|---|---|---|
| **Does the intervention aim to change activity levels?** ***Yes exclusively or substantially / Yes minimally / No*** | **Does the intervention use multiple strategies (three or more)?** ***Yes/No*** | **Is the intervention applied in a single phase?** ***Yes/No*** | **Is the intervention applied for a continued period?** **Yes/No** |
| Yes exclusively/substantially | Yes | Yes | Yes |
| Yes minimally | No | Yes | Yes |
| Yes exclusively/substantially | Yes | No | Yes |
| Yes exclusively/substantially | Yes | Yes | Yes |
| Yes exclusively/substantially | Yes | Yes | Yes |
| Yes exclusively/substantially | No | Yes | Yes |
| Yes exclusively/substantially | Yes | No | Yes |
| Yes exclusively/substantially | No | No | Yes |
| Yes exclusively/substantially | No | No | Yes |
| Yes exclusively/substantially | Yes | Yes | Yes |
| Yes exclusively/substantially | Yes | Yes | Yes |
| No | Yes | Yes | Yes |
| Yes exclusively/substantially | Yes | Yes | Yes |
| Yes exclusively/substantially | Yes | Yes | Yes |
| Yes exclusively/substantially | No | Yes | Yes |
| Yes exclusively/substantially | Yes | No | Yes |
| Yes exclusively/substantially | Yes | No | Yes |
| Yes exclusively/substantially | No | Yes | Yes |
| Yes exclusively/substantially | Yes | Yes | Yes |
| Yes exclusively/substantially | Yes | Yes | Yes |
| Yes exclusively/substantially | Yes | Yes | Yes |
| Yes exclusively/substantially | Yes | Yes | Yes |
| Yes exclusively/substantially | No | Yes | Yes |
| Yes exclusively/substantially | Yes | Yes | Yes |
| Yes exclusively/substantially | No | Yes | Yes |
| Yes exclusively/substantially | No | Yes | Yes |
| Yes exclusively/substantially | No | Yes | Yes |
| Yes exclusively/substantially | No | Yes | Yes |
| Yes exclusively/substantially | Yes | Yes | Yes |
| Yes exclusively/substantially | Yes | Yes | Yes |
| Yes exclusively/substantially | Yes | Yes | Yes |
| No | Yes | Yes | Yes |
| No | Yes | Yes | Yes |
| Yes exclusively/substantially | Yes | Yes | Yes |
| Yes exclusively/substantially | Yes | Yes | Yes |
| No | No | Yes | Yes |

| | | | |
|---|---|---|---|
| Yes exclusively/substantially | Yes | Yes | Yes |
| Yes exclusively/substantially | Yes | Yes | Yes |
| No | Yes | No | Yes |
| Yes exclusively/substantially | Yes | Yes | Yes |
| Yes exclusively/substantially | Yes | Yes | Yes |
| Yes exclusively/substantially | Yes | Yes | Yes |
| No | No | Yes | Yes |
| No | No | Yes | Yes |
| No | Yes | Yes | Yes |
| Yes exclusively/substantially | Yes | Yes | Yes |
| Yes exclusively/substantially | No | Yes | Yes |
| Yes exclusively/substantially | No | Yes | Yes |
| No | No | Yes | Yes |
| Yes exclusively/substantially | Yes | Yes | Yes |
| Yes exclusively/substantially | No | Yes | Yes |
| Yes exclusively/substantially | No | Yes | Yes |
| Yes exclusively/substantially | No | Yes | Yes |
| Yes exclusively/substantially | Yes | Yes | Yes |
| Yes exclusively/substantially | Yes | No | No |
| No | Yes | Yes | Yes |
| Yes exclusively/substantially | No | Yes | Yes |
| Yes exclusively/substantially | Yes | Yes | Yes |
| Yes exclusively/substantially | No | Yes | Yes |
| Yes exclusively/substantially | Yes | Yes | Yes |
| Yes exclusively/substantially | No | Yes | Yes |
| Yes exclusively/substantially | No | Yes | Yes |
| Yes exclusively/substantially | Yes | Yes | Yes |
| Yes exclusively/substantially | Yes | No | Yes |
| No | Yes | Yes | Yes |
| Yes minimally | No | Yes | Yes |
| Yes exclusively/substantially | Yes | Yes | Yes |
| Yes exclusively/substantially | Yes | No | Yes |
| Yes exclusively/substantially | No | Yes | Yes |
| Yes exclusively/substantially | Yes | Yes | Yes |
| Yes exclusively/substantially | Yes | Yes | Yes |
| No | No | Yes | Yes |
| Yes exclusively/substantially | No | No | No |
| Yes exclusively/substantially | Yes | Yes | Yes |
| Yes exclusively/substantially | Yes | Yes | Yes |
| Yes exclusively/substantially | Yes | Yes | Yes |
| Yes exclusively/substantially | Yes | Yes | Yes |
| Yes exclusively/substantially | Yes | Yes | Yes |
| No | No | Yes | Yes |
| Yes exclusively/substantially | Yes | Yes | Yes |
| Yes exclusively/substantially | No | Yes | Yes |
| Yes exclusively/substantially | Yes | Yes | Yes |
| Yes exclusively/substantially | Yes | No | Yes |
| No | No | Yes | Yes |
| Yes exclusively/substantially | Yes | Yes | Yes |
| Yes exclusively/substantially | Yes | No | Yes |

| | | | |
|---|---|---|---|
| Yes exclusively/substantially | Yes | Yes | Yes |
| Yes exclusively/substantially | Yes | Yes | Yes |
| Yes exclusively/substantially | Yes | Yes | Yes |
| Yes exclusively/substantially | Yes | No | Yes |
| Yes minimally | Yes | Yes | Yes |
| Yes exclusively/substantially | Yes | Yes | Yes |
| Yes exclusively/substantially | Yes | Yes | Yes |
| Yes exclusively/substantially | Yes | Yes | Yes |
| No | Yes | Yes | Yes |
| Yes exclusively/substantially | Yes | Yes | Yes |
| No | No | Yes | Yes |
| Yes exclusively/substantially | Yes | Yes | Yes |
| Yes exclusively/substantially | Yes | No | Yes |
| Yes exclusively/substantially | Yes | Yes | Yes |
| Yes exclusively/substantially | Yes | No | No |
| Yes exclusively/substantially | No | Yes | Yes |
| Yes exclusively/substantially | Yes | Yes | Yes |
| Yes minimally | Yes | Yes | Yes |
| Yes exclusively/substantially | No | Yes | Yes |
| Yes exclusively/substantially | No | Yes | Yes |
| Yes exclusively/substantially | No | Yes | Yes |
| Yes exclusively/substantially | Yes | Yes | Yes |
| Yes exclusively/substantially | Yes | No | Yes |
| Yes exclusively/substantially | Yes | Yes | Yes |
| Yes exclusively/substantially | Yes | No | No |
| Yes exclusively/substantially | No | Yes | Yes |
| Yes exclusively/substantially | Yes | Yes | Yes |
| Yes exclusively/substantially | Yes | Yes | Yes |
| Yes exclusively/substantially | No | Yes | Yes |
| No | Yes | No | Yes |
| No | No | Yes | Yes |
| No | No | Yes | Yes |
| Yes exclusively/substantially | No | Yes | Yes |
| Yes exclusively/substantially | No | Yes | Yes |
| Yes exclusively/substantially | Yes | Yes | Yes |
| No | Yes | Yes | Yes |
| Yes exclusively/substantially | Yes | Yes | Yes |
| Yes exclusively/substantially | No | Yes | Yes |
| Yes exclusively/substantially | Yes | Yes | Yes |
| Yes exclusively/substantially | Yes | Yes | Yes |
| Yes exclusively/substantially | Yes | Yes | Yes |
| Yes exclusively/substantially | Yes | Yes | Yes |
| Yes exclusively/substantially | Yes | Yes | Yes |
| Yes exclusively/substantially | Yes | No | Yes |
| Yes exclusively/substantially | Yes | No | Yes |
| No | No | Yes | Yes |
| No | No | Yes | Yes |
| Yes exclusively/substantially | Yes | No | Yes |
| Yes exclusively/substantially | Yes | Yes | Yes |
| Yes exclusively/substantially | No | Yes | Yes |

| | | | |
|---|---|---|---|
| Yes exclusively/substantially | Yes | Yes | Yes |
| Yes exclusively/substantially | No | Yes | Yes |
| Yes minimally | Yes | Yes | Yes |
| Yes exclusively/substantially | Yes | Yes | Yes |
| No | Yes | Yes | Yes |
| No | No | Yes | Yes |
| Yes exclusively/substantially | No | Yes | Yes |
| Yes exclusively/substantially | Yes | No | Yes |
| Yes exclusively/substantially | No | Yes | Yes |
| Yes exclusively/substantially | No | Yes | Yes |
| Yes exclusively/substantially | Yes | No | No |
| Yes exclusively/substantially | Yes | Yes | Yes |
| Yes exclusively/substantially | Yes | Yes | Yes |
| No | No | Yes | Yes |
| No | No | Yes | Yes |
| Yes exclusively/substantially | No | No | Yes |
| Yes exclusively/substantially | Yes | Yes | Yes |
| Yes exclusively/substantially | Yes | Yes | Yes |
| Yes exclusively/substantially | Yes | No | No |
| Yes exclusively/substantially | Yes | No | Yes |
| Yes exclusively/substantially | Yes | Yes | Yes |
| Yes exclusively/substantially | No | Yes | Yes |
| No | No | Yes | Yes |
| Yes exclusively/substantially | Yes | Yes | Yes |
| Yes exclusively/substantially | Yes | Yes | Yes |
| Yes exclusively/substantially | Yes | Yes | Yes |
| Yes exclusively/substantially | Yes | Yes | Yes |
| Yes exclusively/substantially | Yes | Yes | Yes |
| No | Yes | No | Yes |
| No | Yes | Yes | Yes |
| No | Yes | Yes | Yes |
| No | No | Yes | Yes |
| No | No | Yes | Yes |
| No | No | Yes | Yes |
| No | No | Yes | Yes |
| Yes exclusively/substantially | Yes | Yes | Yes |
| Yes exclusively/substantially | Yes | Yes | Yes |
| Yes exclusively/substantially | No | Yes | Yes |
| Yes exclusively/substantially | Yes | Yes | Yes |
| Yes exclusively/substantially | Yes | Yes | Yes |
| Yes exclusively/substantially | No | No | Yes |
| Yes exclusively/substantially | No | No | Yes |
| Yes exclusively/substantially | Yes | Yes | Yes |
| Yes exclusively/substantially | Yes | Yes | Yes |
| Yes exclusively/substantially | Yes | No | Yes |
| Yes exclusively/substantially | No | Yes | Yes |
| Yes exclusively/substantially | No | Yes | Yes |
| Yes exclusively/substantially | No | Yes | Yes |
| Yes exclusively/substantially | No | Yes | Yes |
| Yes exclusively/substantially | Yes | Yes | Yes |

| | | | |
|---|---|---|---|
| Yes exclusively/substantially | No | Yes | Yes |
| Yes exclusively/substantially | Yes | Yes | Yes |
| Yes exclusively/substantially | No | Yes | Yes |
| Yes exclusively/substantially | No | Yes | Yes |
| Yes exclusively/substantially | No | Yes | Yes |
| Yes exclusively/substantially | Yes | Yes | Yes |
| Yes exclusively/substantially | Yes | Yes | Yes |
| Yes exclusively/substantially | No | Yes | Yes |
| Yes exclusively/substantially | Yes | No | Yes |
| Yes exclusively/substantially | Yes | No | Yes |
| Yes exclusively/substantially | Yes | Yes | Yes |
| Yes exclusively/substantially | Yes | Yes | Yes |
| Yes exclusively/substantially | Yes | Yes | Yes |
| Yes exclusively/substantially | Yes | Yes | Yes |
| Yes exclusively/substantially | Yes | Yes | Yes |
| Yes exclusively/substantially | Yes | No | Yes |
| Yes exclusively/substantially | No | Yes | Yes |
| No | Yes | Yes | Yes |
| Yes exclusively/substantially | No | Yes | Yes |
| Yes exclusively/substantially | Yes | No | Yes |
| Yes exclusively/substantially | Yes | No | Yes |
| No | Yes | No | Yes |
| No | Yes | Yes | Yes |
| No | Yes | Yes | Yes |
| Yes exclusively/substantially | Yes | Yes | Yes |
| Yes exclusively/substantially | Yes | Yes | Yes |
| Yes exclusively/substantially | Yes | Yes | Yes |
| Yes exclusively/substantially | No | Yes | Yes |
| Yes exclusively/substantially | Yes | Yes | Yes |
| Yes exclusively/substantially | Yes | Yes | Yes |
| Yes exclusively/substantially | Yes | Yes | Yes |
| No | Yes | Yes | Yes |
| Yes exclusively/substantially | Yes | Yes | Yes |
| Yes exclusively/substantially | Yes | No | Yes |
| Yes exclusively/substantially | Yes | Yes | Yes |
| Yes exclusively/substantially | Yes | Yes | Yes |
| No | Yes | No | Yes |
| Yes exclusively/substantially | No | Yes | Yes |
| Yes exclusively/substantially | No | Yes | Yes |
| Yes exclusively/substantially | No | Yes | Yes |
| Yes exclusively/substantially | No | Yes | Yes |
| Yes exclusively/substantially | No | Yes | Yes |
| Yes exclusively/substantially | No | Yes | Yes |
| Yes exclusively/substantially | No | Yes | Yes |
| Yes exclusively/substantially | No | No | Yes |
| Yes exclusively/substantially | Yes | No | Yes |
| Yes exclusively/substantially | No | No | Yes |
| Yes exclusively/substantially | Yes | No | No |
| No | Yes | No | No |
| Yes exclusively/substantially | Yes | No | No |

| | | | |
|---|---|---|---|
| Yes exclusively/substantially | No | Yes | Yes |
| Yes minimally | No | Yes | Yes |
| Yes minimally | No | Yes | Yes |
| Yes exclusively/substantially | No | Yes | Yes |
| Yes exclusively/substantially | No | Yes | Yes |
| Yes exclusively/substantially | Yes | Yes | Yes |
| Yes exclusively/substantially | No | Yes | Yes |
| Yes exclusively/substantially | No | Yes | Yes |
| Yes exclusively/substantially | Yes | No | Yes |
| Yes exclusively/substantially | Yes | Yes | Yes |
| Yes exclusively/substantially | Yes | Yes | Yes |
| Yes exclusively/substantially | Yes | No | Yes |
| Yes exclusively/substantially | No | Yes | Yes |
| Yes exclusively/substantially | Yes | Yes | Yes |
| Yes exclusively/substantially | Yes | Yes | Yes |
| Yes exclusively/substantially | Yes | Yes | Yes |
| Yes exclusively/substantially | Yes | Yes | Yes |
| Yes exclusively/substantially | No | Yes | Yes |

| Peak intensity and duration | | | Integration |
| --- | --- | --- | --- |
| During how many weeks does the whole intervention last? *Numerical* | For how many weeks does the peak engagement period of intervention last? *Numerical* | What is the level of engagement with the children? *High/Low* | Is the intervention integrated into the normal curriculum/ habits? **Yes completelly/Yes partially/No** |
| 52 | 6 | High | Yes partially |
| 20 | 20 | High | Yes completelly |
| 95.26 | 95.26 | Low | Yes partially |
| 24 | 24 | High | Yes completelly |
| 38.97 | 38.97 | High | Yes completelly |
| 26 | 26 | High | Yes completelly |
| 13 | 4 | High | No |
| 12.99 | 12.99 | High | No |
| 12.99 | 21.65 | High | No |
| 43.3 | 43.3 | High | No |
| 8 | 8 | High | No |
| 23.815 | 10 | High | Yes completelly |
| 38.97 | 38.97 | High | Yes completelly |
| 38.97 | 38.97 | High | Yes completelly |
| 12 | 12 | High | Yes completelly |
| 100.6 | 14 | High | No |
| 100.6 | 14 | High | No |
| 47.63 | 47.63 | Low | No |
| 5 | 5 | Low | Yes partially |
| 30 | 30 | High | Yes completelly |
| 104 | 104 | Low | Yes completelly |
| 104 | 104 | Low | Yes completelly |
| 104 | 104 | Low | No |
| 43.3 | 43.3 | High | Yes completelly |
| 4 | 4 | High | Yes completelly |
| 4 | 4 | High | Yes completelly |
| 52 | 52 | High | Yes completelly |
| 16 | 16 | High | Yes completelly |
| 12 | 12 | Low | No |
| 156 | 32 | High | Yes partially |
| 90.93 | 90.93 | High | Yes partially |
| 12 | 12 | Low | Yes partially |
| 12 | 12 | Low | Yes partially |
| 8 | 8 | High | No |
| 8 | 8 | High | No |
| 8 | 8 | High | No |

| | | | |
|---|---|---|---|
| 12 | 12 | High | Yes completelly |
| 19 | 19 | Low | Yes completelly |
| 104 | 104 | Low | Yes completelly |
| 30.33 | 30.33 | Low | No |
| 156 | 156 | High | Yes completelly |
| 156 | 156 | High | Yes partially |
| 38.97 | 38.97 | Low | Yes completelly |
| 13 | 13 | Low | Yes partially |
| 39 | 39 | Low | Yes completelly |
| 38.97 | 38.97 | Low | Yes partially |
| 22 | 22 | High | Yes completelly |
| 12 | 12 | High | Yes completelly |
| 77.94 | 77.94 | Low | Yes completelly |
| 52 | 52 | High | Yes partially |
| 25.98 | 25.98 | High | Yes completelly |
| 156 | 156 | High | Yes completelly |
| 12 | 12 | High | Yes completelly |
| 8 | 8 | High | Yes partially |
| 18 | 9 | High | No |
| 25 | 25 | Low | Yes partially |
| 12.99 | 12.99 | High | No |
| 103.92 | 25.98 | Low | No |
| 10 | 10 | High | Yes completelly |
| 20 | 20 | High | Yes completelly |
| 104 | 104 | Low | Yes completelly |
| 15 | 15 | High | Yes completelly |
| 104 | 104 | High | Yes partially |
| 52 | 26 | Low | No |
| 12.99 | 12.99 | Low | No |
| 52 | 52 | Low | No |
| 30.31 | 30.31 | Low | No |
| 34.64 | 34.64 | Low | Yes partially |
| 104 | 104 | Low | Yes completelly |
| 9 | 9 | High | No |
| 86.6 | 86.6 | High | Yes completelly |
| 8 | 8 | Low | Yes completelly |
| 38.97 | 26 | Low | No |
| 12.99 | 12.99 | High | Yes completelly |
| 104 | 104 | Low | Yes partially |
| 90.93 | 90.93 | Low | Yes completelly |
| 90.93 | 90.93 | Low | Yes completelly |
| 17.3 | 17.32 | Low | Yes completelly |
| 156 | 156 | Low | Yes completelly |
| 18 | 16 | High | No |
| 18 | 16 | High | No |
| 60.62 | 60.62 | Low | Yes completelly |
| 130 | 130 | High | Yes completelly |
| 24 | 24 | Low | Yes completelly |
| 12.99 | 12.99 | Low | Yes completelly |
| 93.095 | 10 | High | Yes completelly |

| | | | |
|---|---|---|---|
| 20 | 20 | High | Yes partially |
| 90.93 | 90.93 | Low | Yes completelly |
| 43.3 | 43.3 | High | No |
| 52 | 17.32 | Low | No |
| 156 | 156 | Low | Yes completelly |
| 156 | 156 | Low | Yes completelly |
| 156 | 156 | Low | Yes completelly |
| 12 | 12 | Low | Yes completelly |
| 9 | 9 | High | Yes completelly |
| 9 | 9 | High | Yes completelly |
| 47.63 | 47.63 | Low | Yes completelly |
| 104 | 104 | High | Yes partially |
| 30.3 | 30.3 | High | No |
| 30.3 | 30.3 | High | No |
| 51.96 | 17.32 | Low | Yes completelly |
| 52 | 52 | Low | No |
| 10 | 10 | High | Yes completelly |
| 52 | 52 | Low | Yes partially |
| 12 | 12 | High | Yes partially |
| 38.97 | 38.97 | High | Yes completelly |
| 21.65 | 21.65 | Low | Yes completelly |
| 28.145 | 28.145 | Low | Yes partially |
| 100.6 | 14 | High | No |
| 52 | 52 | High | Yes completelly |
| 104 | 16 | Low | Yes completelly |
| 104 | 104 | High | Yes completelly |
| 38.97 | 38.97 | High | Yes partially |
| 38.97 | 38.97 | Low | No |
| 104 | 2 | Low | Yes completelly |
| 7 | 1 | High | No |
| 52 | 52 | Low | Yes partially |
| 1 | 1 | Low | Yes partially |
| 12 | 12 | High | No |
| 25.98 | 25.98 | High | Yes partially |
| 25.98 | 25.98 | High | Yes completelly |
| 104 | 104 | Low | Yes completelly |
| 25.98 | 25.98 | High | Yes completelly |
| 52 | 52 | High | Yes completelly |
| 52 | 52 | Low | Yes partially |
| 46 | 24 | High | Yes completelly |
| 52 | 52 | High | Yes partially |
| 38.97 | 38.97 | Low | Yes partially |
| 104 | 104 | High | Yes completelly |
| 52 | 36 | High | Yes completelly |
| 52 | 16 | High | Yes completelly |
| 11 | 8 | Low | No |
| 11 | 8 | Low | No |
| 104 | 52 | High | Yes partially |
| 208 | 208 | High | Yes completelly |
| 39 | 39 | High | Yes partially |

| | | | |
|---|---|---|---|
| 34.6 | 34.6 | High | No |
| 34.64 | 34.64 | High | No |
| 15 | 15 | High | Yes completelly |
| 52 | 52 | High | Yes completelly |
| 52 | 52 | Low | Yes completelly |
| 12 | 12 | High | Yes completelly |
| 12.99 | 12.99 | Low | No |
| 14 | 7 | High | No |
| 8 | 8 | High | No |
| 208 | 208 | High | Yes completelly |
| 20 | 20 | High | Yes completelly |
| 12 | 12 | High | Yes completelly |
| 12 | 12 | High | Yes completelly |
| 26 | 26 | Low | No |
| 26 | 26 | Low | No |
| 26 | 1 | Low | Yes partially |
| 44 | 44 | High | Yes completelly |
| 44 | 44 | High | Yes completelly |
| 39 | 16 | High | Yes completelly |
| 39 | 16 | High | Yes completelly |
| 12 | 12 | High | No |
| 12 | 12 | Low | No |
| 12.99 | 12.99 | Low | Yes completelly |
| 12 | 12 | High | No |
| 12 | 12 | High | No |
| 26 | 26 | Low | Yes partially |
| 26 | 26 | Low | Yes partially |
| 10 | 10 | High | No |
| 26 | 26 | Low | No |
| 34.64 | 34.64 | Low | No |
| 34.64 | 34.64 | Low | No |
| 35.62 | 35.62 | Low | No |
| 35.62 | 35.62 | Low | No |
| 35.62 | 35.62 | Low | No |
| 35.62 | 35.62 | Low | No |
| 39 | 39 | High | Yes completelly |
| 30.31 | 30.31 | Low | No |
| 30.31 | 30.31 | Low | No |
| 16 | 16 | High | Yes completelly |
| 17 | 17 | High | Yes partially |
| 3 | 3 | High | No |
| 3 | 3 | High | No |
| 43.3 | 43.3 | High | Yes partially |
| 9 | 9 | High | Yes completelly |
| 24 | 12 | High | No |
| 1 | 1 | Low | Yes completelly |
| 17.3 | 17.3 | Low | No |
| 25.98 | 25.98 | Low | No |
| 25.98 | 25.98 | Low | No |
| 12.99 | 12.99 | High | No |

| | | | |
|---|---|---|---|
| 12.99 | 12.99 | Low | No |
| 12.99 | 12.99 | High | Yes completelly |
| 12.99 | 12.99 | Low | Yes completelly |
| 13 | 13 | Low | Yes completelly |
| 25.98 | 25.98 | Low | Yes partially |
| 17.32 | 17.32 | Low | Yes completelly |
| 104 | 104 | Low | Yes partially |
| 104 | 104 | High | Yes completelly |
| 104 | 52 | High | Yes partially |
| 104 | 104 | High | No |
| 47.63 | 47.63 | Low | Yes completelly |
| 73.7 | 73.7 | High | Yes partially |
| 130 | 130 | Low | Yes completelly |
| 130 | 130 | High | Yes completelly |
| 130 | 130 | High | Yes completelly |
| 43.3 | 43.3 | High | No |
| 10 | 10 | High | Yes completelly |
| 52 | 9 | High | No |
| 1 | 1 | Low | Yes completelly |
| 30.31 | 30.31 | Low | Yes partially |
| 52 | 12 | Low | No |
| 43 | 43 | High | No |
| 6 | 6 | High | No |
| 30.31 | 30.31 | Low | Yes completelly |
| 43.3 | 43.3 | Low | Yes partially |
| 77.94 | 77.94 | High | Yes partially |
| 208 | 208 | Low | Yes partially |
| 24 | 24 | High | Yes completelly |
| 6 | 6 | High | Yes completelly |
| 24 | 24 | High | Yes partially |
| 43.3 | 43.3 | High | Yes partially |
| 52 | 4 | High | No |
| 52 | 4 | High | No |
| 77.94 | 12 | High | No |
| 12 | 12 | High | Yes completelly |
| 44 | 44 | High | Yes completelly |
| 38.97 | 19.485 | High | Yes partially |
| 79.74 | 79.74 | Low | Yes partially |
| 43.3 | 43.3 | High | Yes completelly |
| 75 | 75 | High | Yes completelly |
| 31 | 31 | Low | Yes completelly |
| 31 | 31 | Low | Yes completelly |
| 26 | 26 | High | Yes partially |
| 12 | 12 | High | No |
| 12 | 12 | High | No |
| 12 | 12 | High | Yes partially |
| 12 | 12 | High | Yes partially |
| 26 | 14 | High | Yes partially |
| 26 | 14 | High | Yes completelly |
| 26 | 14 | High | Yes partially |

| | | | |
|---|---|---|---|
| 12 | 12 | High | Yes completelly |
| 20 | 20 | High | Yes completelly |
| 25 | 25 | High | Yes completelly |
| 24 | 24 | High | No |
| 43.3 | 43.3 | High | Yes completelly |
| 43.3 | 43.3 | Low | Yes partially |
| 34.64 | 34.64 | Low | Yes completelly |
| 95.26 | 86.6 | Low | Yes completelly |
| 104 | 12 | Low | No |
| 24 | 24 | High | Yes completelly |
| 24 | 24 | High | Yes completelly |
| 24 | 24 | High | No |
| 4 | 4 | High | Yes completelly |
| 121.24 | 121.24 | High | Yes completelly |
| 121.24 | 121.24 | High | Yes completelly |
| 34.64 | 34.64 | Low | Yes completelly |
| 52 | 52 | High | Yes completelly |
| 156 | 156 | High | Yes partially |

| Flexibility | Choice | Fun factor | |
|---|---|---|---|
| Is the intervention designed to be implemented in a flexible manner/tailored to specific participants? *Yes/No* | Is choice of activity/diet designed into the intervention? Yes/No | How enticing do you find this strategy? Boring/Neutral/Fun | How enticing do you think children in the intended age group would find this strategy? *Boring/Neutral/Fun* |
| Yes | No | Fun | Fun |
| No | No | Neutral | Fun |
| No | No | Fun | Fun |
| No | No | Fun | Fun |
| No | No | Fun | Fun |
| No | Yes | Neutral | Fun |
| No | No | Fun | Fun |
| No | No | Fun | Fun |
| No | No | Fun | Fun |
| Yes | Yes | Fun | Fun |
| No | No | Fun | Fun |
| Yes | No | Neutral | Fun |
| Yes | No | Boring | Fun |
| Yes | No | Boring | Boring |
| No | No | Boring | Boring |
| No | No | Neutral | Fun |
| No | No | Neutral | Fun |
| Yes | No | Fun | Fun |
| No | No | Fun | Fun |
| No | No | Fun | Fun |
| No | No | Neutral | Neutral |
| No | No | Fun | Fun |
| Yes | No | Boring | Boring |
| No | No | Neutral | Boring |
| No | No | Neutral | Fun |
| No | No | Neutral | Fun |
| Yes | Yes | Fun | Fun |
| Yes | No | Boring | Boring |
| No | No | Fun | Fun |
| No | No | Fun | Fun |
| No | No | Neutral | Neutral |
| Yes | Yes | Fun | Fun |
| Yes | Yes | Fun | Fun |
| No | No | Neutral | Boring |
| Yes | No | Fun | Boring |
| No | No | Fun | Boring |

| | | | |
|---|---|---|---|
| No | No | Fun | Fun |
| No | Yes | Boring | Boring |
| Yes | No | Fun | Fun |
| No | No | Boring | Fun |
| No | No | Fun | Fun |
| No | No | Fun | Fun |
| No | No | Fun | Fun |
| Yes | Yes | Fun | Fun |
| No | No | Fun | Fun |
| Yes | No | Fun | Neutral |
| No | No | Fun | Neutral |
| No | No | Fun | Fun |
| No | No | Boring | Fun |
| Yes | Yes | Fun | Fun |
| No | No | Boring | Neutral |
| Yes | No | Fun | Fun |
| Yes | No | Fun | Fun |
| No | Yes | Boring | Boring |
| No | No | Fun | Fun |
| No | Yes | Neutral | Neutral |
| Yes | No | Neutral | Boring |
| Yes | No | Boring | Boring |
| Yes | No | Neutral | Boring |
| No | No | Boring | Boring |
| Yes | No | Neutral | Fun |
| No | No | Fun | Fun |
| Yes | No | Boring | Neutral |
| No | No | Neutral | Neutral |
| No | No | Neutral | Neutral |
| No | No | Fun | Fun |
| No | No | Fun | Fun |
| Yes | Yes | Fun | Fun |
| Yes | No | Boring | Boring |
| No | Yes | Neutral | Fun |
| Yes | No | Neutral | Fun |
| Yes | No | Fun | Fun |
| No | Yes | Fun | Fun |
| Yes | No | Boring | Fun |
| No | No | Fun | Fun |
| Yes | Yes | Fun | Fun |
| Yes | Yes | Boring | Boring |
| Yes | No | Boring | Boring |
| No | No | Neutral | Boring |
| No | No | Boring | Fun |
| No | No | Fun | Boring |
| Yes | No | Fun | Fun |
| Yes | No | Fun | Fun |
| No | No | Boring | Boring |
| No | Yes | Fun | Fun |
| Yes | Yes | Fun | Neutral |

| | | | |
|---|---|---|---|
| No | Yes | Fun | Fun |
| No | No | Boring | Boring |
| Yes | Yes | Fun | Fun |
| No | No | Fun | Fun |
| No | No | Boring | Neutral |
| No | No | Fun | Neutral |
| No | No | Fun | Neutral |
| Yes | No | Fun | Fun |
| No | No | Fun | Fun |
| No | No | Fun | Fun |
| No | No | Fun | Fun |
| Yes | Yes | Fun | Fun |
| Yes | No | Boring | Boring |
| No | No | Boring | Neutral |
| No | No | Boring | Boring |
| Yes | No | Boring | Boring |
| Yes | Yes | Fun | Fun |
| No | No | Neutral | Neutral |
| Yes | No | Fun | Fun |
| No | No | Fun | Fun |
| No | No | Neutral | Boring |
| No | No | Neutral | Fun |
| No | No | Neutral | Fun |
| No | No | Fun | Boring |
| No | Yes | Neutral | Neutral |
| No | Yes | Fun | Fun |
| No | No | Fun | Fun |
| Yes | No | Neutral | Fun |
| Yes | Yes | Boring | Boring |
| No | No | Fun | Fun |
| No | Yes | Neutral | Boring |
| No | No | Neutral | Boring |
| No | Yes | Fun | Fun |
| No | No | Neutral | Fun |
| No | Yes | Boring | Boring |
| No | Yes | Boring | Boring |
| No | No | Fun | Fun |
| Yes | No | Fun | Fun |
| Yes | Yes | Neutral | Fun |
| Yes | No | Fun | Fun |
| Yes | Yes | Boring | Boring |
| Yes | No | Neutral | Boring |
| No | No | Fun | Fun |
| No | Yes | Fun | Fun |
| Yes | Yes | Boring | Fun |
| No | Yes | Boring | Boring |
| No | Yes | Boring | Boring |
| Yes | No | Neutral | Fun |
| No | No | Neutral | Boring |
| No | No | Fun | Fun |

| | | | |
|---|---|---|---|
| No | No | Fun | Fun |
| Yes | No | Fun | Fun |
| No | No | Fun | Fun |
| No | No | Boring | Fun |
| No | No | Fun | Neutral |
| No | No | Boring | Boring |
| No | No | Fun | Fun |
| No | Yes | Neutral | Fun |
| No | Yes | Fun | Fun |
| No | No | Fun | Fun |
| No | No | Fun | Fun |
| No | No | Fun | Fun |
| No | No | Fun | Fun |
| Yes | No | Neutral | Boring |
| No | No | Neutral | Boring |
| No | No | Boring | Boring |
| No | No | Fun | Fun |
| No | No | Fun | Fun |
| No | No | Fun | Fun |
| No | No | Fun | Fun |
| Yes | Yes | Fun | Fun |
| Yes | Yes | Fun | Fun |
| Yes | Yes | Fun | Neutral |
| Yes | Yes | Boring | Boring |
| No | No | Boring | Neutral |
| Yes | No | Neutral | Neutral |
| Yes | No | Neutral | Neutral |
| No | No | Fun | Fun |
| Yes | No | Boring | Boring |
| Yes | No | Boring | Boring |
| Yes | No | Boring | Boring |
| No | No | Boring | Boring |
| No | No | Fun | Boring |
| No | No | Fun | Boring |
| No | No | Fun | Boring |
| Yes | Yes | Fun | Fun |
| No | Yes | Fun | Fun |
| No | No | Fun | Fun |
| No | No | Fun | Fun |
| No | No | Fun | Boring |
| No | Yes | Fun | Fun |
| No | Yes | Fun | Fun |
| No | No | Neutral | Fun |
| No | No | Fun | Fun |
| Yes | Yes | Fun | Fun |
| Yes | No | Boring | Boring |
| Yes | No | Boring | Boring |
| No | Yes | Fun | Fun |
| No | No | Boring | Neutral |
| No | Yes | Fun | Fun |

| | | | |
|---|---|---|---|
| No | No | Boring | Boring |
| Yes | Yes | Fun | Fun |
| No | Yes | Boring | Boring |
| No | No | Neutral | Boring |
| Yes | No | Fun | Boring |
| Yes | No | Fun | Fun |
| Yes | No | Boring | Fun |
| No | No | Neutral | Fun |
| Yes | No | Boring | Neutral |
| Yes | No | Boring | Neutral |
| Yes | No | Neutral | Boring |
| Yes | No | Fun | Neutral |
| No | Yes | Boring | Fun |
| Yes | Yes | Boring | Fun |
| Yes | Yes | Boring | Fun |
| No | No | Fun | Fun |
| Yes | Yes | Fun | Boring |
| Yes | Yes | Fun | Fun |
| No | No | Fun | Boring |
| No | No | Neutral | Boring |
| Yes | No | Boring | Boring |
| Yes | Yes | Fun | Fun |
| No | No | Fun | Fun |
| No | No | Boring | Boring |
| No | No | Fun | Fun |
| No | No | Fun | Fun |
| Yes | Yes | Fun | Fun |
| No | Yes | Fun | Fun |
| No | Yes | Boring | Boring |
| No | No | Neutral | Neutral |
| No | Yes | Fun | Boring |
| No | No | Boring | Boring |
| No | No | Neutral | Neutral |
| Yes | Yes | Fun | Boring |
| Yes | Yes | Fun | Fun |
| No | No | Fun | Fun |
| No | No | Fun | Fun |
| Yes | Yes | Fun | Fun |
| Yes | No | Fun | Fun |
| No | No | Neutral | Boring |
| No | No | Boring | Boring |
| No | No | Boring | Boring |
| No | No | Boring | Boring |
| No | No | Fun | Fun |
| No | No | Fun | Fun |
| No | No | Fun | Fun |
| No | No | Fun | Fun |
| No | No | Fun | Fun |
| No | No | Fun | Fun |
| No | No | Fun | Fun |

| | | | |
|---|---|---|---|
| No | No | Fun | Fun |
| No | No | Neutral | Neutral |
| No | No | Fun | Fun |
| No | Yes | Fun | Fun |
| No | No | Fun | Neutral |
| No | No | Fun | Fun |
| No | No | Fun | Fun |
| No | Yes | Fun | Fun |
| No | Yes | Fun | Fun |
| No | No | Fun | Fun |
| No | No | Fun | Fun |
| Yes | Yes | Neutral | Neutral |
| No | No | Fun | Fun |
| No | No | Fun | Fun |
| Yes | No | Neutral | Boring |
| No | No | Fun | Fun |
| No | No | Fun | Fun |
| No | No | Fun | Fun |

| Resonance | Mechanism of action and recipient | | |
|---|---|---|---|
| Is the intervention experienced by children via someone external or unusual? *Yes/No* | Does the intervention have an explicit component that requires the child to participate? *Yes/No* | Does the intervention have an explicit component of education/information provision for the child? *Yes/No* | Does the intervention have an explicit component aiming to change the social environment of the child? *Yes/No* |
| Yes | Yes | Yes | Yes |
| No | No | Yes | No |
| Yes | Yes | Yes | Yes |
| No | Yes | Yes | Yes |
| No | Yes | Yes | Yes |
| Yes | Yes | Yes | No |
| Yes | Yes | Yes | Yes |
| No | No | Yes | No |
| No | No | Yes | No |
| Yes | Yes | No | No |
| Yes | Yes | Yes | Yes |
| No | No | Yes | Yes |
| No | Yes | No | Yes |
| No | Yes | Yes | Yes |
| No | Yes | Yes | No |
| Yes | Yes | Yes | No |
| Yes | No | No | Yes |
| Yes | Yes | Yes | No |
| No | No | Yes | Yes |
| Yes | Yes | Yes | Yes |
| Yes | No | Yes | No |
| No | No | No | No |
| Yes | No | Yes | No |
| No | Yes | Yes | Yes |
| Yes | No | Yes | No |
| Yes | No | Yes | No |
| No | Yes | No | Yes |
| Yes | No | Yes | Yes |
| Yes | Yes | Yes | No |
| No | Yes | Yes | Yes |
| Yes | Yes | Yes | Yes |
| Yes | No | Yes | Yes |
| Yes | No | Yes | Yes |
| Yes | Yes | Yes | Yes |
| No | No | Yes | Yes |
| No | No | Yes | Yes |

| | | | |
|---|---|---|---|
| Yes | Yes | Yes | Yes |
| Yes | Yes | No | Yes |
| No | Yes | Yes | Yes |
| Yes | Yes | Yes | Yes |
| Yes | Yes | Yes | Yes |
| Yes | Yes | Yes | Yes |
| Yes | No | Yes | No |
| No | Yes | Yes | No |
| Yes | Yes | Yes | Yes |
| Yes | Yes | No | Yes |
| Yes | Yes | No | No |
| Yes | Yes | Yes | No |
| No | Yes | No | No |
| Yes | Yes | Yes | Yes |
| No | Yes | No | No |
| No | Yes | No | No |
| No | Yes | No | No |
| No | Yes | Yes | Yes |
| Yes | Yes | Yes | Yes |
| Yes | No | Yes | Yes |
| Yes | Yes | No | No |
| No | No | No | Yes |
| No | No | Yes | No |
| No | Yes | Yes | Yes |
| No | No | No | Yes |
| Yes | Yes | No | No |
| No | No | Yes | Yes |
| Yes | Yes | Yes | Yes |
| Yes | Yes | Yes | Yes |
| Yes | Yes | Yes | Yes |
| Yes | Yes | Yes | Yes |
| No | Yes | Yes | Yes |
| No | No | Yes | Yes |
| Yes | Yes | Yes | Yes |
| No | Yes | Yes | Yes |
| Yes | No | Yes | No |
| Yes | Yes | Yes | Yes |
| Yes | Yes | Yes | Yes |
| No | Yes | Yes | Yes |
| No | Yes | Yes | Yes |
| No | Yes | Yes | No |
| No | No | Yes | Yes |
| No | No | No | No |
| Yes | Yes | Yes | Yes |
| Yes | No | No | Yes |
| No | No | No | Yes |
| Yes | Yes | Yes | Yes |
| No | No | No | Yes |
| Yes | No | No | Yes |
| No | Yes | No | Yes |

| | | | |
|---|---|---|---|
| No | Yes | Yes | Yes |
| No | No | Yes | Yes |
| Yes | Yes | No | No |
| Yes | Yes | Yes | Yes |
| No | No | No | Yes |
| No | No | No | Yes |
| No | No | No | Yes |
| No | Yes | Yes | Yes |
| No | Yes | Yes | No |
| Yes | Yes | Yes | No |
| Yes | No | Yes | No |
| Yes | Yes | Yes | Yes |
| Yes | Yes | No | Yes |
| Yes | No | Yes | Yes |
| No | No | Yes | Yes |
| Yes | No | Yes | Yes |
| No | Yes | Yes | No |
| No | No | Yes | Yes |
| Yes | Yes | No | No |
| Yes | Yes | No | No |
| No | Yes | Yes | No |
| No | Yes | Yes | Yes |
| Yes | Yes | Yes | Yes |
| No | Yes | Yes | Yes |
| No | Yes | Yes | Yes |
| No | No | Yes | No |
| Yes | Yes | No | Yes |
| Yes | Yes | Yes | Yes |
| No | No | Yes | No |
| Yes | Yes | Yes | No |
| Yes | Yes | Yes | No |
| Yes | No | Yes | No |
| Yes | Yes | No | No |
| Yes | Yes | No | No |
| Yes | Yes | Yes | Yes |
| Yes | No | Yes | No |
| Yes | Yes | Yes | Yes |
| No | Yes | Yes | Yes |
| Yes | Yes | Yes | Yes |
| Yes | Yes | Yes | Yes |
| No | Yes | Yes | Yes |
| No | Yes | Yes | Yes |
| No | No | Yes | Yes |
| Yes | No | Yes | Yes |
| No | Yes | Yes | Yes |
| Yes | Yes | No | No |
| Yes | Yes | No | No |
| No | Yes | Yes | Yes |
| No | Yes | No | Yes |
| Yes | Yes | No | Yes |

| | | | |
|---|---|---|---|
| Yes | Yes | No | Yes |
| Yes | Yes | No | No |
| No | No | Yes | Yes |
| No | Yes | Yes | Yes |
| No | No | Yes | Yes |
| No | No | Yes | Yes |
| Yes | Yes | No | Yes |
| No | Yes | No | Yes |
| Yes | Yes | Yes | Yes |
| No | Yes | No | No |
| Yes | Yes | No | No |
| No | No | Yes | No |
| No | No | Yes | No |
| Yes | No | No | Yes |
| Yes | No | Yes | Yes |
| No | No | Yes | Yes |
| Yes | Yes | Yes | Yes |
| Yes | Yes | Yes | Yes |
| Yes | Yes | Yes | Yes |
| Yes | Yes | Yes | Yes |
| No | Yes | No | Yes |
| No | Yes | No | Yes |
| No | Yes | No | Yes |
| No | No | Yes | No |
| No | No | Yes | No |
| No | No | Yes | Yes |
| No | No | Yes | Yes |
| Yes | Yes | Yes | Yes |
| No | No | Yes | Yes |
| Yes | No | Yes | Yes |
| Yes | No | Yes | Yes |
| Yes | No | Yes | Yes |
| Yes | No | Yes | Yes |
| Yes | No | Yes | Yes |
| Yes | No | Yes | Yes |
| No | Yes | Yes | Yes |
| Yes | Yes | Yes | Yes |
| No | No | Yes | Yes |
| Yes | Yes | Yes | Yes |
| Yes | Yes | Yes | No |
| No | No | Yes | No |
| Yes | No | Yes | No |
| Yes | Yes | Yes | Yes |
| Yes | Yes | Yes | Yes |
| Yes | Yes | Yes | Yes |
| Yes | No | Yes | No |
| Yes | No | Yes | No |
| No | No | Yes | Yes |
| No | No | Yes | Yes |
| Yes | Yes | Yes | Yes |

| | | | |
|---|---|---|---|
| Yes | No | Yes | Yes |
| Yes | Yes | Yes | Yes |
| Yes | No | Yes | Yes |
| Yes | Yes | No | Yes |
| No | No | Yes | No |
| No | Yes | Yes | Yes |
| Yes | Yes | Yes | Yes |
| No | Yes | No | No |
| Yes | Yes | Yes | Yes |
| Yes | Yes | Yes | No |
| Yes | Yes | Yes | No |
| No | Yes | Yes | Yes |
| No | No | Yes | Yes |
| No | Yes | Yes | Yes |
| No | Yes | Yes | Yes |
| No | Yes | Yes | Yes |
| Yes | Yes | No | Yes |
| Yes | Yes | Yes | Yes |
| No | No | No | Yes |
| Yes | Yes | Yes | Yes |
| No | No | No | Yes |
| Yes | Yes | Yes | Yes |
| Yes | Yes | No | No |
| Yes | No | Yes | Yes |
| No | Yes | Yes | Yes |
| No | Yes | Yes | Yes |
| No | No | Yes | Yes |
| Yes | Yes | No | No |
| No | Yes | Yes | Yes |
| Yes | Yes | Yes | Yes |
| No | Yes | Yes | Yes |
| No | Yes | Yes | Yes |
| No | Yes | Yes | Yes |
| Yes | Yes | Yes | Yes |
| Yes | Yes | Yes | Yes |
| No | Yes | No | Yes |
| Yes | Yes | Yes | Yes |
| No | Yes | No | Yes |
| No | Yes | No | No |
| Yes | Yes | No | No |
| No | Yes | No | No |
| No | Yes | No | No |
| Yes | Yes | No | No |
| Yes | Yes | Yes | Yes |
| Yes | Yes | Yes | Yes |
| Yes | Yes | Yes | Yes |
| Yes | Yes | Yes | Yes |
| Yes | Yes | Yes | Yes |
| Yes | Yes | Yes | Yes |
| Yes | Yes | Yes | Yes |

| | | | |
|---|---|---|---|
| Yes | Yes | No | No |
| No | No | Yes | No |
| No | No | Yes | No |
| Yes | Yes | No | No |
| No | Yes | Yes | No |
| No | No | Yes | Yes |
| No | Yes | No | No |
| No | Yes | No | No |
| Yes | Yes | Yes | Yes |
| No | No | Yes | Yes |
| No | No | Yes | Yes |
| No | Yes | Yes | Yes |
| Yes | Yes | Yes | No |
| No | Yes | No | No |
| No | Yes | Yes | No |
| No | Yes | Yes | Yes |
| No | Yes | Yes | Yes |
| No | Yes | Yes | No |

| | Commercial interests |
|---|---|
| **Does the intervention have an explicit component aiming to change the physical environment of the child?** ***Yes/No*** | **Are commercial interests involved in the trial/intervention?** ***Yes/No*** |
| No | No |
| No | No |
| Yes | No |
| Yes | No |
| Yes | No |
| No | No |
| No | No |
| No | Yes |
| No | No |
| No | No |
| No | No |
| No | No |
| No | No |
| No | No |
| No | No |
| No | No |
| No | No |
| No | No |
| Yes | No |
| No | No |
| No | No |
| Yes | No |
| No | No |
| No | No |
| No | No |
| No | No |
| Yes | No |
| No | No |
| No | No |
| Yes | No |
| Yes | No |
| No | No |
| No | No |
| No | No |
| No | No |
| No | No |

| | |
|---|---|
| No | No |
| Yes | No |
| Yes | No |
| Yes | No |
| Yes | No |
| Yes | No |
| No | No |
| Yes | Yes |
| Yes | Yes |
| No | No |
| Yes | No |
| No | No |
| No | No |
| No | No |
| No | No |
| No | No |
| No | No |
| No | No |
| No | No |
| Yes | No |
| No | No |
| Yes | No |
| No | No |
| No | No |
| Yes | No |
| No | No |
| Yes | No |
| Yes | No |
| No | No |
| No | No |
| No | No |
| Yes | No |
| No | No |
| No | No |
| Yes | No |
| No | No |
| Yes | No |
| Yes | No |
| Yes | Yes |
| Yes | No |
| Yes | No |
| No | No |
| Yes | No |
| No | No |
| No | No |
| Yes | No |
| Yes | Yes |
| No | Yes |
| No | No |
| Yes | No |

| | |
|---|---|
| No | No |
| Yes | No |
| No | No |
| No | No |
| No | No |
| Yes | No |
| Yes | No |
| No | No |
| No | No |
| No | No |
| No | Yes |
| No | No |
| Yes | No |
| No | No |
| No | No |
| No | Yes |
| Yes | No |
| Yes | No |
| No | Yes |
| No | No |
| No | No |
| No | No |
| No | No |
| No | No |
| No | No |
| Yes | Yes |
| Yes | No |
| No | No |
| No | No |
| No | No |
| No | No |
| No | No |
| No | No |
| No | No |
| No | No |
| Yes | No |
| Yes | No |
| No | No |
| Yes | Yes |
| No | No |
| Yes | No |
| No | No |
| No | No |
| No | No |
| No | No |
| No | No |
| No | No |
| Yes | No |
| Yes | No |
| No | No |

| | |
|---|---|
| Yes | No |
| No | No |
| No | No |
| No | No |
| Yes | No |
| No | No |
| No | No |
| No | No |
| Yes | No |
| No | Yes |
| Yes | No |
| No | No |
| No | No |
| No | No |
| No | No |
| No | No |
| No | No |
| No | No |
| No | No |
| No | No |
| No | Yes |
| No | Yes |
| Yes | Yes |
| No | No |
| No | No |
| No | No |
| No | No |
| Yes | No |
| Yes | No |
| No | Yes |
| No | Yes |
| No | Yes |
| No | Yes |
| No | Yes |
| No | Yes |
| No | No |
| No | No |
| No | No |
| No | No |
| No | No |
| No | No |
| No | No |
| Yes | Yes |
| No | No |
| No | No |
| No | No |
| No | No |
| No | No |
| No | No |
| Yes | No |

| | |
|---|---|
| No | No |
| Yes | No |
| No | No |
| No | Yes |
| No | No |
| No | No |
| Yes | No |
| No | No |
| Yes | No |
| Yes | No |
| Yes | No |
| No | Yes |
| Yes | No |
| Yes | No |
| Yes | No |
| No | No |
| No | No |
| Yes | No |
| No | No |
| No | No |
| No | No |
| Yes | No |
| Yes | No |
| No | No |
| Yes | No |
| No | No |
| Yes | No |
| Yes | Yes |
| Yes | No |
| No | No |
| No | No |
| No | No |
| No | No |
| No | No |
| No | No |
| Yes | No |
| No | No |
| No | No |
| No | No |
| No | No |
| No | No |
| No | No |
| No | No |
| No | No |
| No | No |
| No | No |
| No | No |
| Yes | No |
| Yes | No |
| No | No |

| | |
|---|---|
| No | No |
| No | No |
| No | No |
| No | No |
| No | No |
| Yes | No |
| No | No |
| Yes | Yes |
| No | No |
| No | No |
| No | No |
| No | No |
| No | No |
| Yes | No |
| Yes | No |
| Yes | Yes |
| Yes | No |
| No | No |
